# Smoking is associated with worse outcomes of COVID-19 particularly among younger adults: A systematic review and meta-analysis

**DOI:** 10.1101/2020.09.22.20199802

**Authors:** Roengrudee Patanavanich, Stanton A. Glantz

**Affiliations:** Center for Tobacco Control Research and Education, Department of Medicine, University of California San Francisco San Francisco, CA, U.S.A. 94143-1390; Department of Community Medicine, Faculty of Medicine Ramathibodi Hospital, Mahidol University, Thailand

## Abstract

**Background:** Smoking impairs lung immune functions and damages upper airways, increasing risks of contracting and severity of infectious diseases.

**Methods:** We searched PubMed and Embase for studies published from January 1-May 25, 2020. We included studies reporting smoking behavior of COVID-19 patients and progression of disease, including death. We used a random effects meta-analysis and used meta-regression and lowess regressions to examine relationships in the data.

**Results:** We identified 47 peer-reviewed papers with a total of 31,871 COVID-19 patients, 5,759 (18.1%) experienced disease progression and 5,734 (18.0%) with a history of smoking. Among smokers, 29.2% experienced disease progression, compared with 21.1% of non-smokers. The meta-analysis confirmed an association between smoking and COVID-19 progression (OR 1.56, 95% CI 1.32-1.83, p=0.001). Smoking was associated with increased risk of death from COVID-19 (OR 1.19, 95% CI 1.05-1.34, p=0.007). We found no significant difference (p=0.432) between the effects of smoking on COVID-19 disease progression between adjusted and unadjusted analyses, suggesting that smoking is an independent risk factor for COVID-19 disease progression. We also found the risk of having COVID-19 progression among younger adults (p=0.023), with the effect most pronounced among people under about 45 years old.

**Conclusions:** Smoking is an independent risk for having severe progression of COVID-19, including mortality. The effects seem to be higher among young people. Smoking prevention and cessation should remain a priority for the public, physicians, and public health professionals during the COVID-19 pandemic.

## BACKGROUND

Coronavirus disease 2019 (COVID-19) first occurred in Wuhan, China in December 2019 and has spread rapidly around the world. As of June 2020, the virus had infected over 6 million people and caused more than 300,000 deaths. Old people and those with pre-existing medical conditions including respiratory disease, hypertension, diabetes, cardiovascular disease, and cancer are more vulnerable to becoming critically ill when infected [1].

Smoking appears to enhance the risk of COVID-19 by its biological effects and behaviors of smokers. Smoking impairs lung function and pulmonary immune function, compromising the body’s defense mechanisms against infections [2]. Smoking is also a well-established risk factor for chronic diseases that are linked to more severe COVID-19. The World Health Organization (WHO) has advised the public that smoking could increase the risk of contracting COVID-19 because the behavior of smokers involves contact of fingers with the lips and removal of the protective face masks to smoke [3].

Our recent meta-analysis of the 19 peer-reviewed papers found that smokers have double the odds of COVID-19 progression risk [4]. Some people argue that the association between underlying health conditions and risk factors such as smoking to the severity of COVID-19 is still unclear due to inadequate adjustment of confounding factors [5]. In addition, it is unclear whether the association between smoking and severity of COVID-19 varies by age. This paper updates and extends our previous meta-analysis [4] of 19 studies to add 28 additional studies, including 7 that provided adjusted odds ratios and compared the association between smoking and COVID-19 disease progression between unadjusted analyses with adjusted analyses to examine whether smoking is an independent risk factor. We also assessed the effect of age of patients and conducted a sub-analysis for the risk of smoking on the mortality of COVID-19.

## METHODS

This study followed the Preferred Reporting in Systematic Reviews and Meta-Analyses (PRISMA) guidelines and is registered with PROSPERO (CRD42020186864).

### Data source and search strateg*y*

We conducted a systematic search using PubMed and Embase on May 25, 2020, with the search term: “((smoking) OR (characteristics) OR (risk factors) OR (retrospective*) OR (outcomes) OR (smoker*)) AND ((COVID-19) OR (COVID) OR (coronavirus) OR (sars cov-2) OR (sars cov 2))” for studies published between January 1, 2020 and May 25, 2020. A total of 2,600 studies were retrieved through PubMed and 1,962 studies through Embase.

### Eligibility criteria

Eligible studies included published peer-reviewed observational studies, retrospective cohort studies, prospective cohort studies, cross-sectional studies, case series, and case reports that reported demographic characteristics, comorbidities specifically smoking status, clinical manifestations, and clinical or disease outcomes of COVID-19 patients on disease progression of COVID-19 to more severe or critical conditions or death. We included both inpatient and outpatient settings. We excluded studies that did not report smoking status and outcomes, studies of children, studies that included other coronavirus infection and not specifically to COVID-19, studies that the number of smokers was zero or omitted, and studies in which all patients had the same outcome. There were no language restrictions.

### Study Selection and Data Extraction

One author (RP) extracted information for each study, screened the abstract or the full text, with questions resolved through discussion among both authors (Figure A1).

The exposure group for our analysis were those who had a history of smoking (current smokers or former smokers) and unexposed group was never smokers, non-smoker, or not having a smoking history. Outcomes were progression of COVID-19 to more severe or critical conditions or death. Definitions of smoking status and disease progression for each study are shown in Table A1.

### Quality Assessment

We evaluated the quality of studies using a modification of the ACROBAT-NRSI[6] tool on 5 domains: study population, exposure measurement, outcome assessment, measurement of confounders, and adequate follow-up. Each one of these domains was scored from 0 (low risk of bias) to 2 (high risk of bias) and the average score of each study was computed and discussed among both authors (Additional file and Table A2). Studies with the average score higher than 1 were considered high risk and excluded in a sensitivity analysis.

### Statistical analyses

Our meta-analyses were based on unadjusted odds ratios (OR) that were either reported in the studies or computed unadjusted OR and 95% confidence interval (CI) using the number of smokers (current and former) and never smokers with and without disease progression. We also did a sensitivity analysis to determine the results changed when the 5 studies with high risk of bias were excluded.

We performed subgroup analyses of (1) the studies that reported association of smoking on COVID-19 mortality and (2) the association of COVID-19 disease progression between current smokers and never smokers (i.e., excluding former smokers) using the studies that reported whether the patient was a current, former, or never smoker (as separate categories).

We also computed the pooled adjusted OR using the studies that reported adjusted OR and 95% CI and compared it with the pooled unadjusted OR.

The results of the included studies were performed with random-effect models using the Stata version 14.0 *metan* command and *metabias* command with Egger’s test for the presence of publication bias. We used *metareg* command (with dummy variables to account for the pairing of adjusted and unadjusted ORs) to determine whether the adjustment of OR affected the results. To examine the effects by age, we used *lowess* command to generate a nonparametric fit estimate of the relationship between odds of disease progression and mean age of each study. We also tested for a trend using *metareg* command with mean age of each study as a continuous variable.

## RESULTS

### Study characteristics

From the total of 4,562 studies we found from our search, 237 studies were considered retrospective cohorts, prospective cohorts, or case series that provided clinical and demographic characteristics of COVID-19 patients. From the 237 studies, 83 studies reported smoking status of the patients, but only 47 studies [7-53] reported smoking status and disease progression of COVID-19 that met our inclusion and exclusion criteria (Figure A1).

Of the 47 studies (Table S1), 33 [10-13, 17-21, 23, 26-29, 31, 35, 37-53] were from China, 8 [8, 9, 14, 16, 22, 25, 33, 34] from the US, 3 [15, 32, 36] from Italy, 1 [7] from the UK, 1 [24] from South Korea, and 1 [30] from 11 countries in Asia, Europe, and North America (China, Japan, South Korea, Turkey, Spain, Italy, Germany, France, UK, Canada, and US).

Eight studies [9, 11, 17, 26, 30, 34, 36, 49] assessed whether the patient was a current, former, or never smoker (as separate categories), 15 [12, 14, 16, 20, 31, 39-43, 45, 46, 50, 51, 53] studies assessed whether the patient was a “current smoker”, 24 [7, 8, 10, 13, 15, 18, 19, 21-25, 27-29, 32, 33, 35, 37, 38, 44, 47, 48, 52] studies assessed whether the patient had a “history of smoking” (current or former).

Clinical outcome was defined as death in 8 studies [11, 15, 25, 30, 32, 33, 44, 53], intensive care unit (ICU) admission or requirement of mechanical ventilation in 6 studies [7, 9, 14, 16, 20, 22], prolonged viral shredding in 2 studies [42, 43], severe or critical (respiratory distress with respiratory rate ≥ 30/min, or oxygen saturation ≤ 93% at rest, or oxygenation index ≤ 300 mmHg, based on the diagnostic and treatment guideline for SARS-CoV-2 issued by Chinese National Health Committee [54] or the American Thoracic Society guidelines [55] for community acquired pneumonia) in 18 studies, the primary composite end point (ICU admission, the use of mechanical ventilation, or death) in 2 studies [17, 46], abnormal chest imaging in 1 study [51], acute cardiac injury in 1 study [18], progression of disease to more severe status (including increasing oxygen supplement, pneumonia exacerbation, transferred to ICU, and sepsis) in 10 studies [12, 19, 23, 24, 29, 31, 33, 36, 37, 47].

There were 5 studies [7, 12, 14, 32, 38] with high risk of bias scores (Table S2).

### Smoking and COVID-19 disease progression

A total of 31,871 COVID-19 patients are included in our meta-analysis, 5,759 of whom (18.1%) experienced disease progression and 5,734 (18.0%) with a history of smoking. Among smokers, 29.2% experienced disease progression, compared with 21.1% of non-smokers. The meta-analysis showed an association between smoking and COVID-19 progression (OR 1.56, 95% CI 1.32-1.83, p=0.001) (Figure 1). There was statistically significant moderate heterogeneity among the studies (I^2^=45.3%, p=0.001) and no evidence of publication bias (p=0.243).

**Figure 1.**
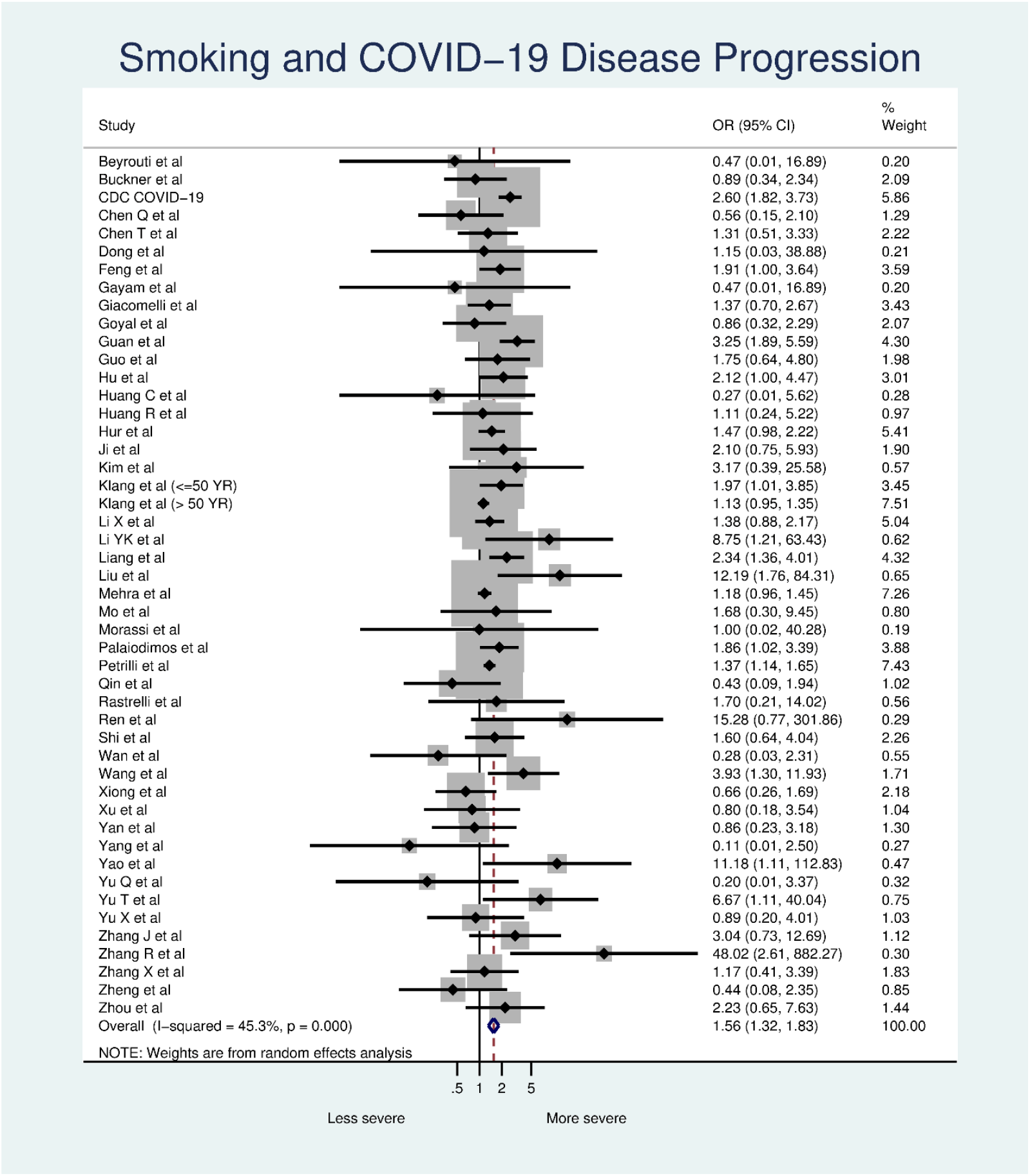
Smoking and COVID-19 disease progression.

Smoking prevalence was significantly higher among patients with disease progression than those without disease progression (p=0.04 by paired t-test).

### Smoking and COVID-19 mortality

Eight studies [11, 15, 25, 30, 32, 33, 44, 53] (n = 13,272) reported death as the outcome of COVID-19 patients. The meta-analysis showed an association between smoking and COVID-19 mortality (OR 1.19, 95% CI 1.05-1.34, p=0.007; Figure S3). There was no evidence of heterogeneity among the studies (I^2^=0%, p=0.654) and no evidence of publication bias (p=0.632).

### Studies of current smokers vs. never smokers

The 8 studies [9, 11, 17, 26, 30, 34, 36, 49] that reported current smokers vs. never smokers yielded an elevated point estimate for the effect of smoking on COVID-19 progression (OR 1.42, 95% CI 0.96-2.1, p=0.079; Figure A4), but it did not reach conventional statistical significance. There was statistically significant moderate heterogeneity (I^2^=58.6%, p=0.018) and no evidence of publication bias (p=0.390).

### Unadjusted vs. Adjusted analyses

Seven studies [19, 25, 29, 30, 33, 34, 47] reported adjusted ORs. The point estimate for the adjusted ORs (OR 1.84, 95% CI 1.17-2.90, p=0.008; Figure A5, top) was higher than the point estimate for unadjusted ORs (OR 1.60, 95% CI 1.17-2.20, p=0.003; Figure A5, bottom), but this difference was not significant (p=0.432). For the adjusted ORs, the heterogeneity among the studies was high and statistically significant (I^2^=80.9%, p=0.001) with an evidence of publication bias (p=0.031). For the unadjusted ORs, the heterogeneity among the studies was moderate and statistically significant (I^2^=67.5%, p=0.003) with an evidence of publication bias (p=0.016).

### Association between smoking and COVID-19 disease progression by age

The odds of COVID-19 disease progression between smokers and non-smokers dropped as the patients’ mean age increased across studies, with the drop most pronounced for studies where the mean age was less than about 45 years old (Figure 2). A meta-regression of the odds of COVID-19 disease progression between smokers and non-smokers and the patients’ mean age showed that each the odds of disease progression dropped statistically significantly by a factor of 0.82 (95% CI 0.69-0.97, p =0.023) per 10 years.

**Figure 2.**
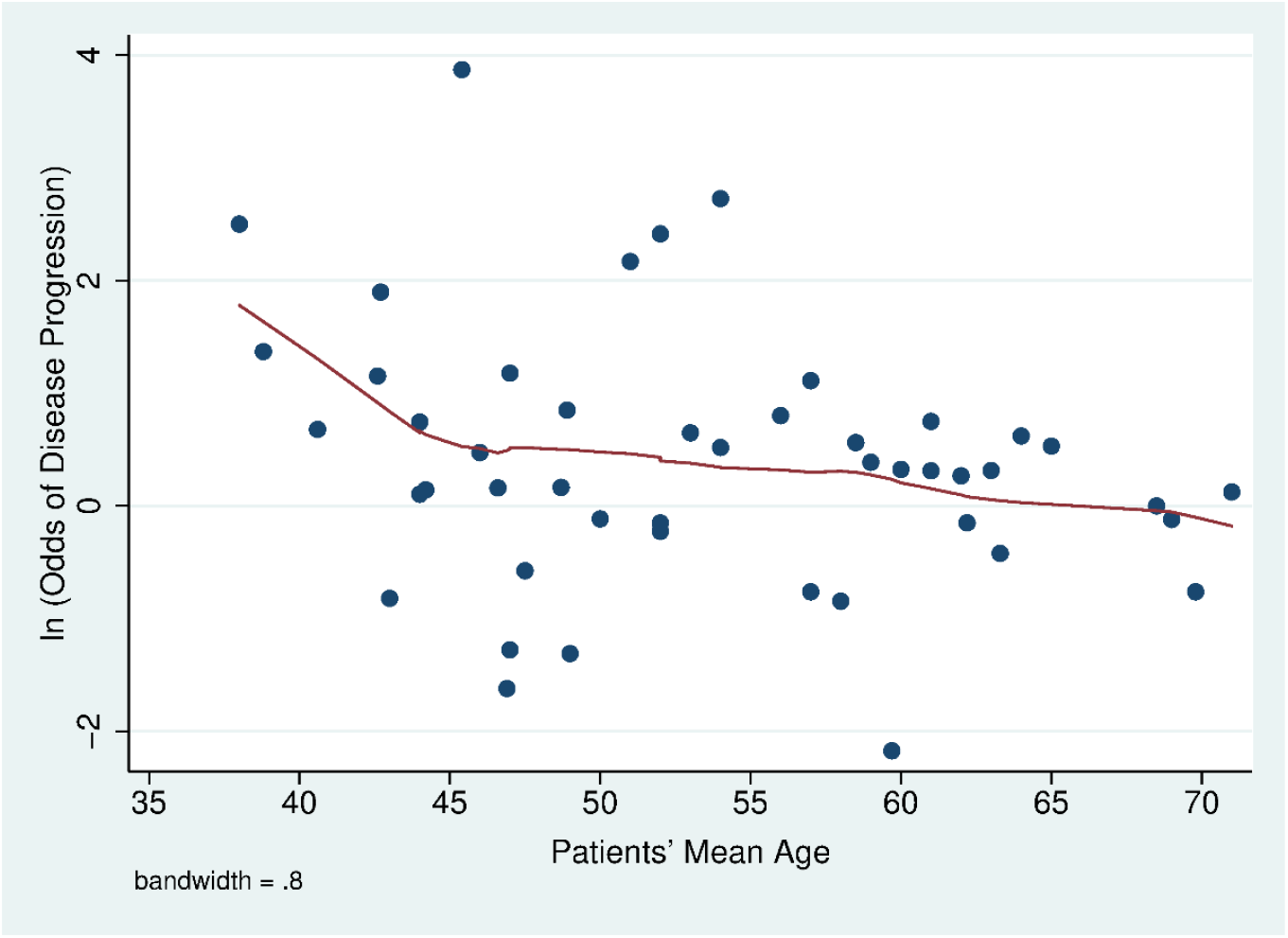
As the mean age of patients in a study falls the odds of COVID-19 progression increases. Solid line fit using a lowess regession.

### Sensitivity analysis

Dropping the 5 studies [7, 12, 14, 32, 38] with high risk of bias scores had little effect on the odds of COVID-19 disease progression (OR 1.57, 95% CI 1.32-1.86, p=0.001; Figure S2). The heterogeneity among the studies was moderate and statistically significant (I^2^=50.6%, p=0.001) and there was no evidence of publication bias (p=0.169).

## DISCUSSION

With more than twice as many studies available compared to our earlier meta-analysis [4], smoking remains a risk factor for COVID-19 disease progression, with smokers having 1.56 times the odds of progression in COVID-19 severity than non-smokers (Figure 1). The risk of smoking on COVID-19 disease progression was not changed significantly by adjusting for confounders, which suggests that smoking is an independent risk of COVID-19 progression. We also find that smokers are at increased risk of death from COVID-19 (Figure A3). These findings are not surprising because the well-established evidence that smoking is associated with a higher risk of viral infection [2]. In the past pandemics such as influenza [2] and Cov-MERS [56] smoking is also among leading risk factors for worse outcomes.

Young smokers appear to have a higher risk of COVID-19 disease progression than older smokers (Figure 2). A recent study also found that young adults are more medically vulnerable to severe COVID-19 illness if they are smokers [57]. The greater effect of smoking among young people is particularly important because in the U.S., almost 40% of COVID-19 patients are aged 18-44 years [58], and in China, 44% of COVID-19 patients are adults aged 20-49 years [59]. Even so, younger adults tend to perceiving lesser infection-fatality risks of COVID-19 [60] so that they are less likely to protect themselves from the infection. Our finding is consistent with a recent meta-analysis study [61] which concluded that age was negatively significantly associated with the effect of smoking on COVID-19 disease severity.

While there is not yet direct peer reviewed evidence of the effect of e-cigarette use on COVID-19 risk, the fact that e-cigarettes have similar adverse effects on pulmonary immune function [62] combined with the fact that e-cigarette use is concentrated among younger people, raises concerns and points to the need to collect data on e-cigarette use and COVID-19 risk.

Some have argued that smoking has a protective effect against COVID-19 because of the low prevalence of reported among COVID patients [63-65]. This is not new. There was also rumors that smoking protected patients from developing Cov-SARS during the 2003 pandemic [66]. However, a case-control study of 447 patients showed that smoking did not protect patients from contracting Cov-SARS after adjusting for confounding by age, gender, contact history, and occupation [66].

Reported smoking prevalence in the 33 studies in China ranged from 1.4% to 29.8% (median = 7.3%), which was substantially lower than 27.7% (52.1% for men and 2.7% for women) smoking prevalence in 2015 [67]. Four studies [9, 14, 16, 34] in the U.S. that reported the smoking prevalence among current smokers ranged from 1.3% to 33.3% (median=5.2%), which was also lower than 13.7% (15.6% for men and 12.0% for women) smoking prevalence in 2018 [68]. The other 4 studies [8, 22, 25, 33] in the U.S. reported the ever-smoking prevalence ranged from 13.3%-33.5%, which was also lower than 41.9% (47.2% for men and 37.3% for women) in 2017 [69]. One study [36] in Italy reported the smoking prevalence among current smokers of 3.2%, which was also substantial lower than 21.1% (26% for men and 17.2% for women) in 2016 [70]. The remaining studies that reported the ever-smoking prevalence (2 studies[15, 32] in Italy, 1 study [7] in UK and 1 study [24] in South Korea) were also lower than the countries’ rates (Italy: 16.7%-30% vs.43.9% (50% for men and 38.3% for women) in 2010 [71]; UK: 16.7% vs. 40.2 (44.3% for men and 36.5% for women) in 2018 [72]; South Korea: 18.5% vs.39.1% (81.6% for men and 6.9% for women) in 2015 [73]). These low levels of reported smoking among COVID-19 patients may reflect the difficulty of obtaining accurate smoking histories among seriously ill patients, especially when most medical facilities are operating at or above normal capacity. Despite the fact that the reported levels of smoking have been below population prevalences; however, the reported smoking prevalence among people with worse outcomes was significantly higher than those with less severe outcomes (13.5% vs. 9.7%, p=0.042).

## Limitations

The studies used a variety of clinical definitions of disease progression and smoking status (Table A1). The varying definitions of disease progression include severity of disease based on clinical manifestations, development to more severe conditions, increasing oxygen supplements, prolonged viral shredding, organ injuries, ICU admission, and death. These varying definitions likely introduced increased variance in the results.

Moreover, smoking was significantly associated with death – a clearly defined endpoint – in the 8 studies [11, 15, 25, 30, 32, 33, 44, 53] that used this endpoint.

Most studies reported smoking status as having a smoking history which was unclear how they categorized former smokers. Of the 47 studies we reviewed, only 8 [9, 11, 17, 26, 30, 34, 36, 49] reported all three smoking categories (current, former, and never smokers). A meta-analysis of these studies found that current smoking was associated with a similar increase in the point estimate for the odds of disease progression (OR 1.42, 95% CI 0.96-2.1; Figure A4) as the other studies (OR 1.49, 95% CI 1.21-1.84), but the odds for current smoking did not reach conventional statistical significance (p=0.079).

Studies that only describe patient smoking history as “smoking history” or “history of smoking” do not provide enough information to analyze smoking as a risk factor given the fact that time since quitting could have significant influence on the patient’s outcomes.

All these limitations add to misclassification errors, which tend to bias results toward the null, suggest that this analysis underestimates the risk of smoking in terms of increasing COVID-19 severity.

The effects of smoking on COVID-19 disease progression by age reported in our paper is limited to the mean age of the studies. Individual level data on smoking, age, e-cigarette use, demographics and other risk factors are needed to perform a more sophisticated analysis. In addition, most of the studies were retrospective cohorts or case series, there might be recall bias, and could not conclude a causal relationship. Most of the meta-analyses in this study had moderate and statistically significant heterogeneity; the reliability of the meta-analysis might be compromised.

## Conclusions

Smoking is an independent risk associated with severe progression of COVID-19, including mortality. The effects seem to be prominent among younger adults. Smoking prevention and cessation should remain a priority for the public, physicians, and public health professionals during the COVID-19 pandemic.

## Data Availability

All data used to prepare this paper are available from the cited sources.

## Abbreviations

COVID-19: Coronavirus disease 2019
CI: Confidence Interval
OR: Odds ratios
ICU: Intensive care unit

## Acknowledgement

Not applicable

## Authors’ contributions

RP developed the idea for the study, collected, analyzed the data, and wrote the first draft of the manuscript. SAG assisted with revising and refining the manuscript.

## Availability of data and materials

All data used to prepare this paper are available from the cited sources.

## Ethics approval and consent to participate

Not applicable

## Consent for publication

Not applicable

## ADDITIONAL FILE

### Risk of Bias Assessment

We evaluated the quality of studies using a modification of the ACROBAT-NRSI[6] tool on 5 domains: study population, exposure measurement, outcome assessment, measurement of confounders, and adequate follow-up. Each one of these domains was scored 0 for low risk of bias, 1 for moderate risk of bias, and 2 for high risk of bias and the average score of each study was computed and discussed among both authors. Studies with the average score higher than 1 were considered high risk and excluded in a sensitivity analysis.

#### Study population

- Low risk: studies included at least 50 patients
- Moderate risk: studies included at least 20 patients
- High risk: studies included less than 20 patients

#### Exposure measurement

- Low risk: studies reported 3 categories of smoking status: current, former, and never smokers
- Moderate risk: studies reported 2 categories of smoking status
- High risk: studies reported only 1 category of smoking status

#### Outcome assessment

- Low risk: studies reported a clear definition of outcomes
- High risk: studies did not report specific outcomes

#### Measurement of confounders

- Low risk: adjustment for confounders
- High risk: unadjusted analyses

#### Adequate follow-up

- Low risk: identify duration of data collection
- High risk: duration of data collection is unclear

#### References

References are the same as cited in the main text.

**Table A1.**
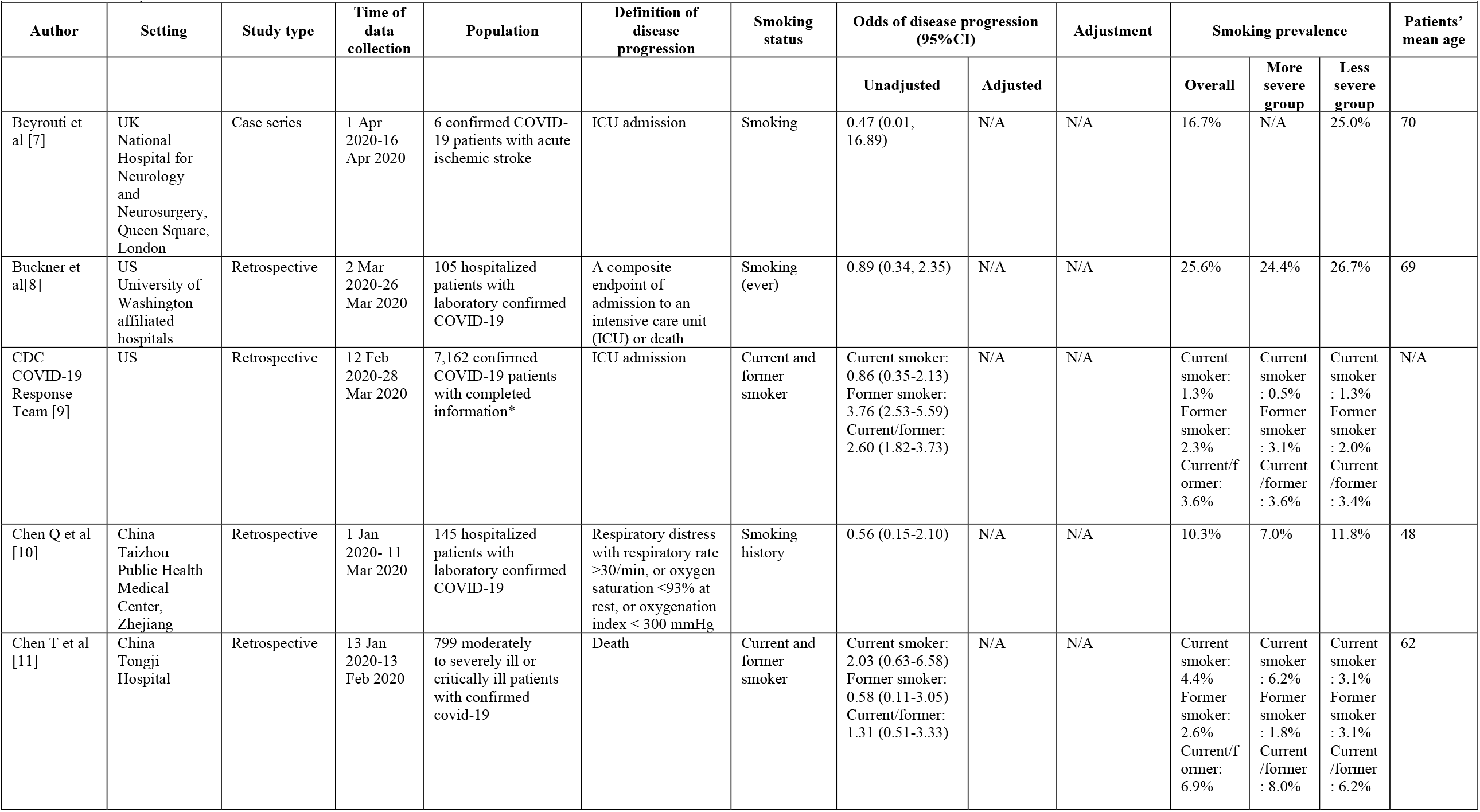
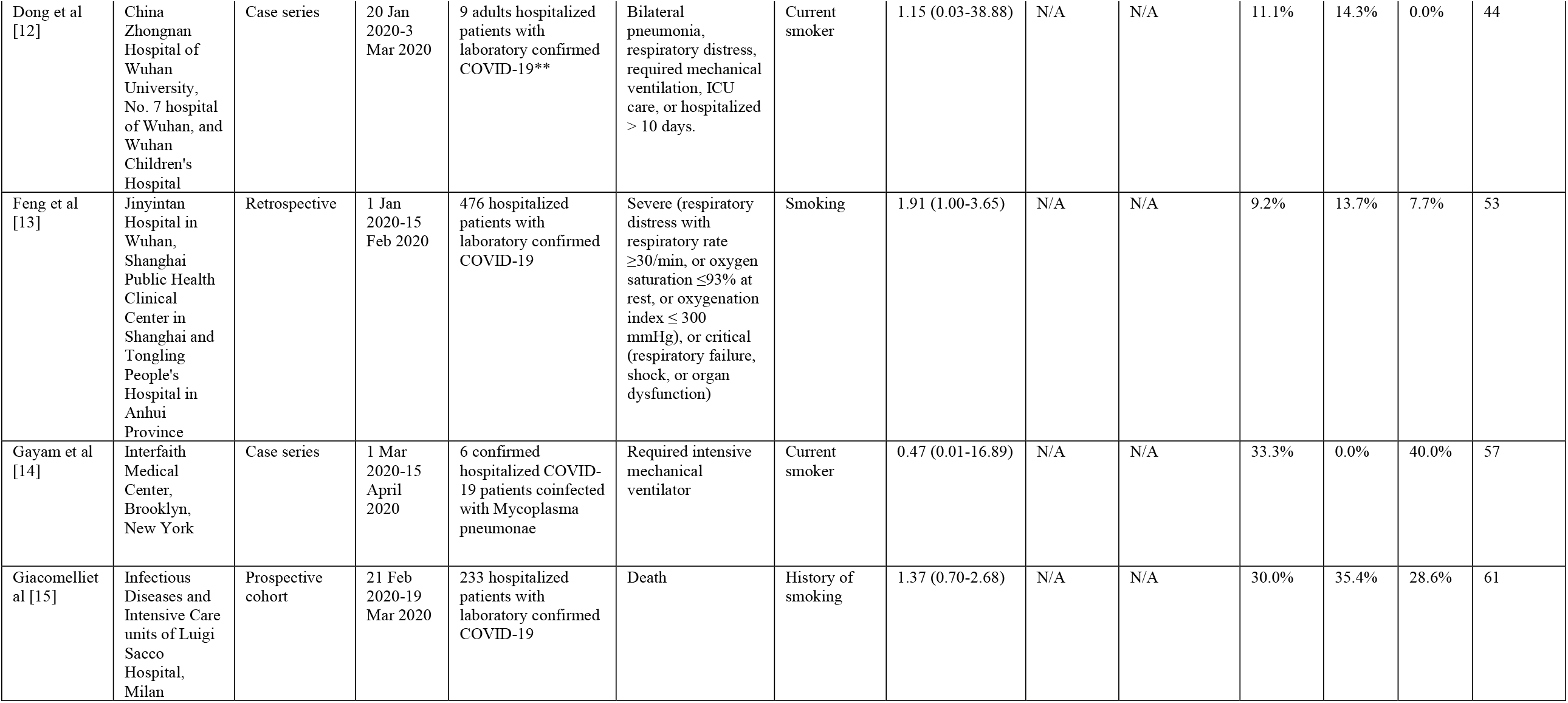
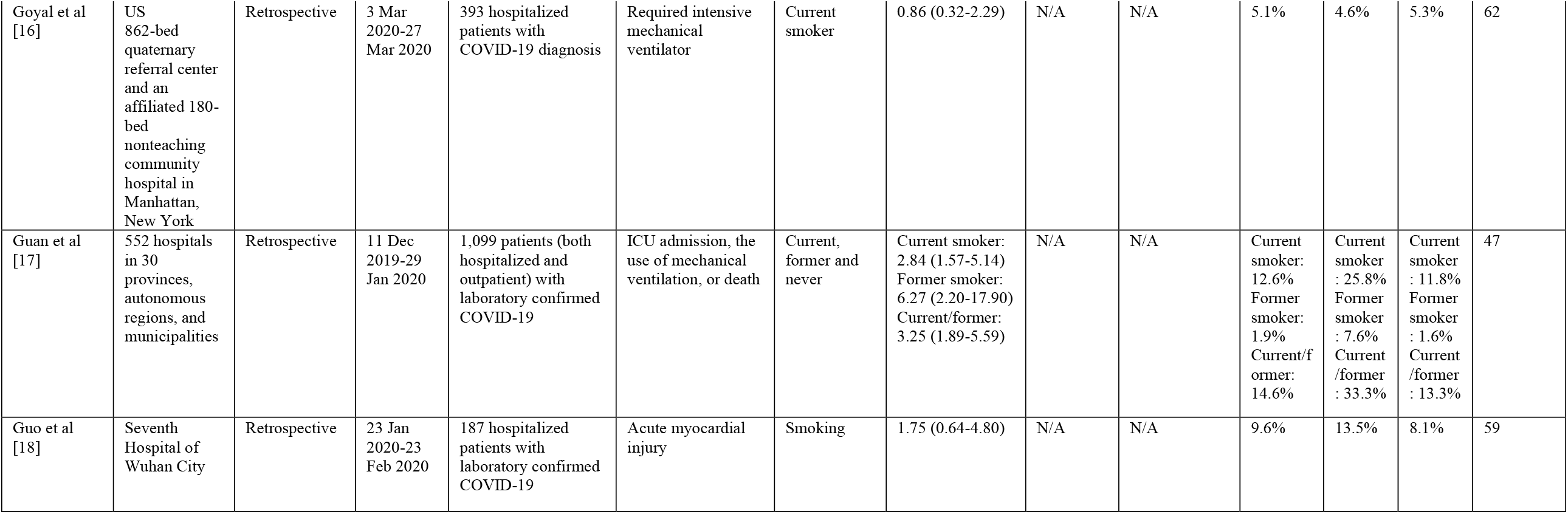
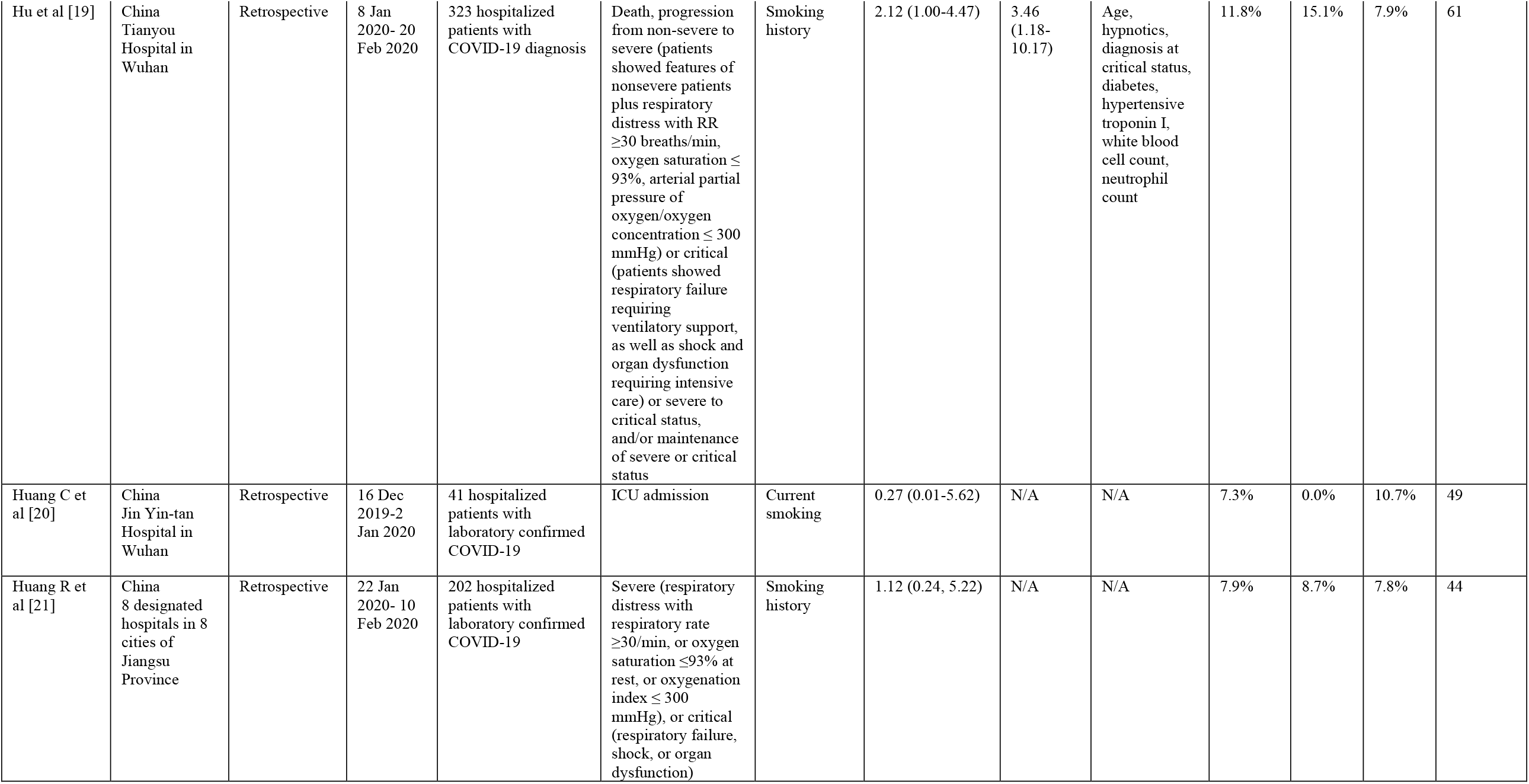
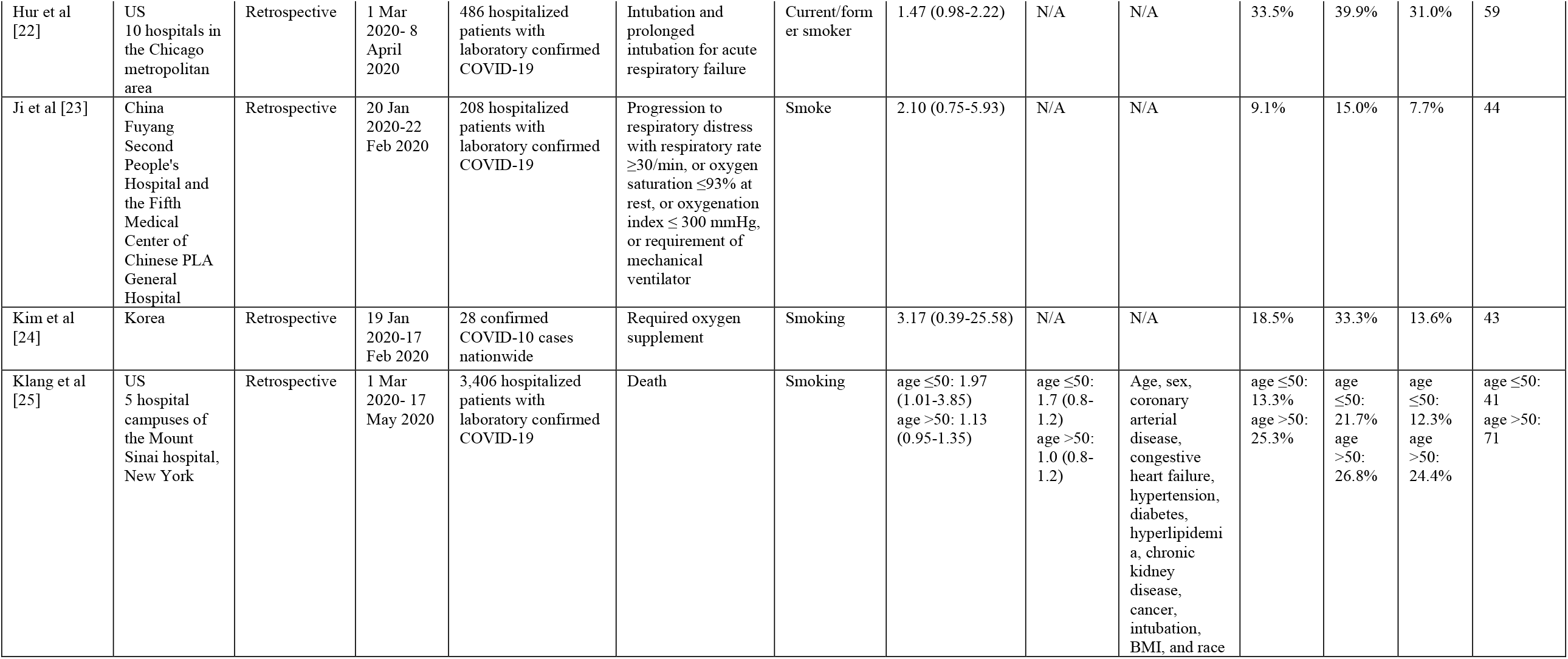
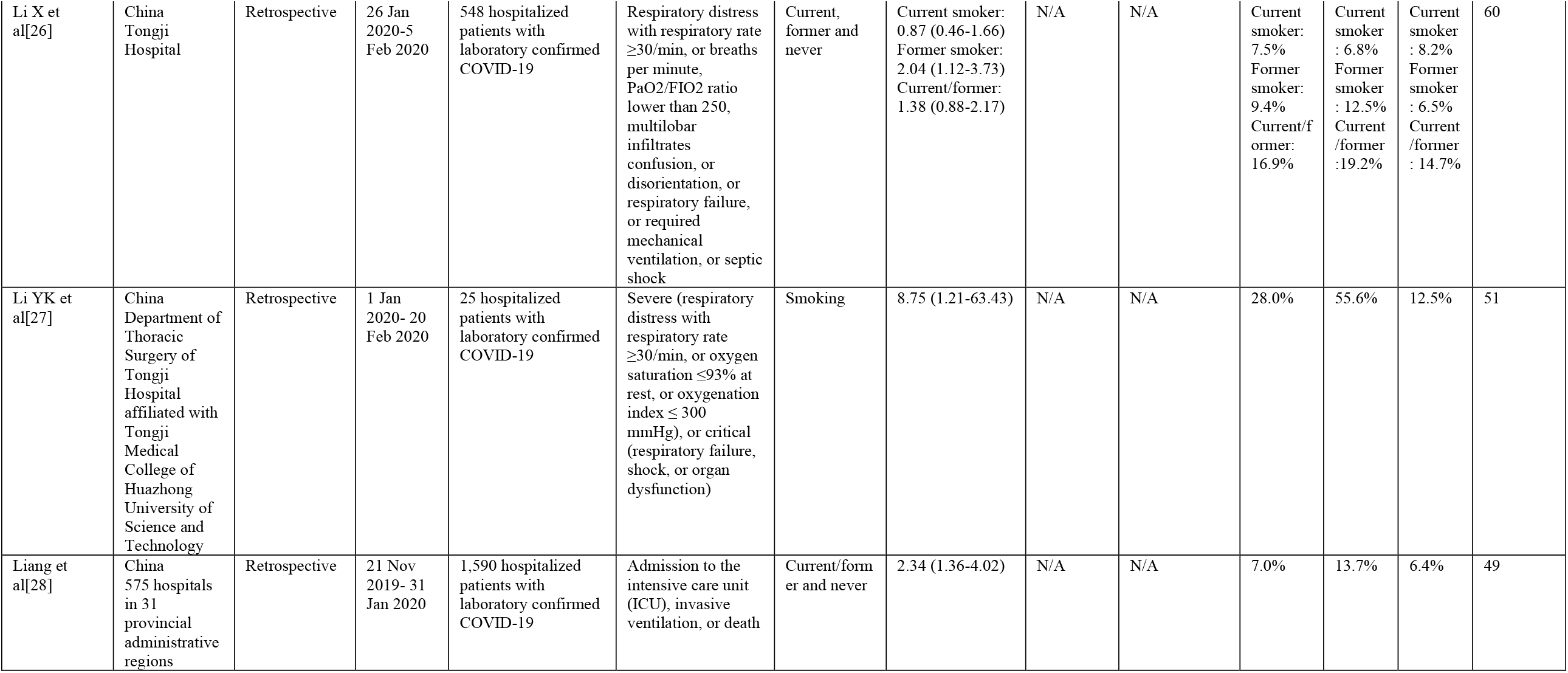
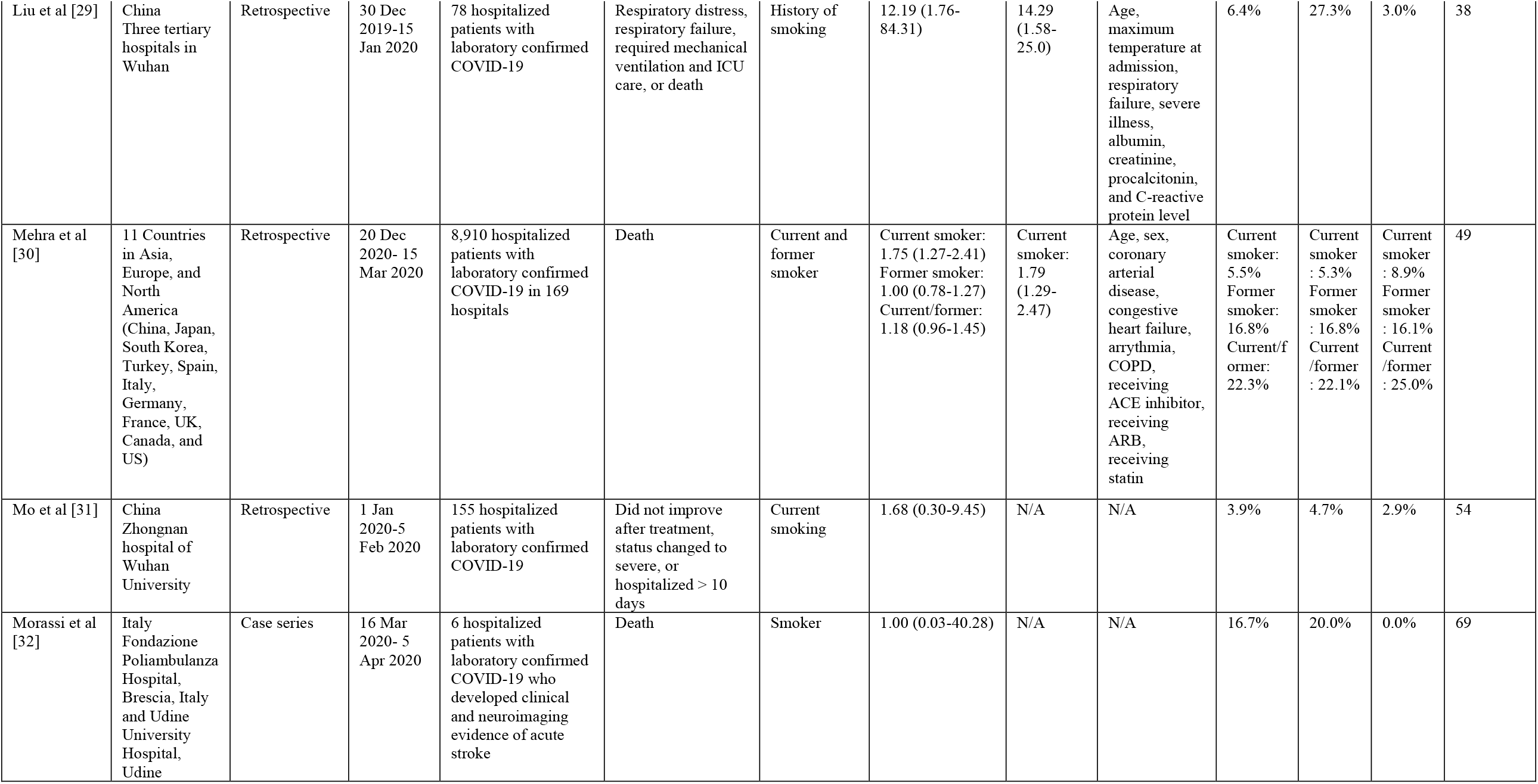
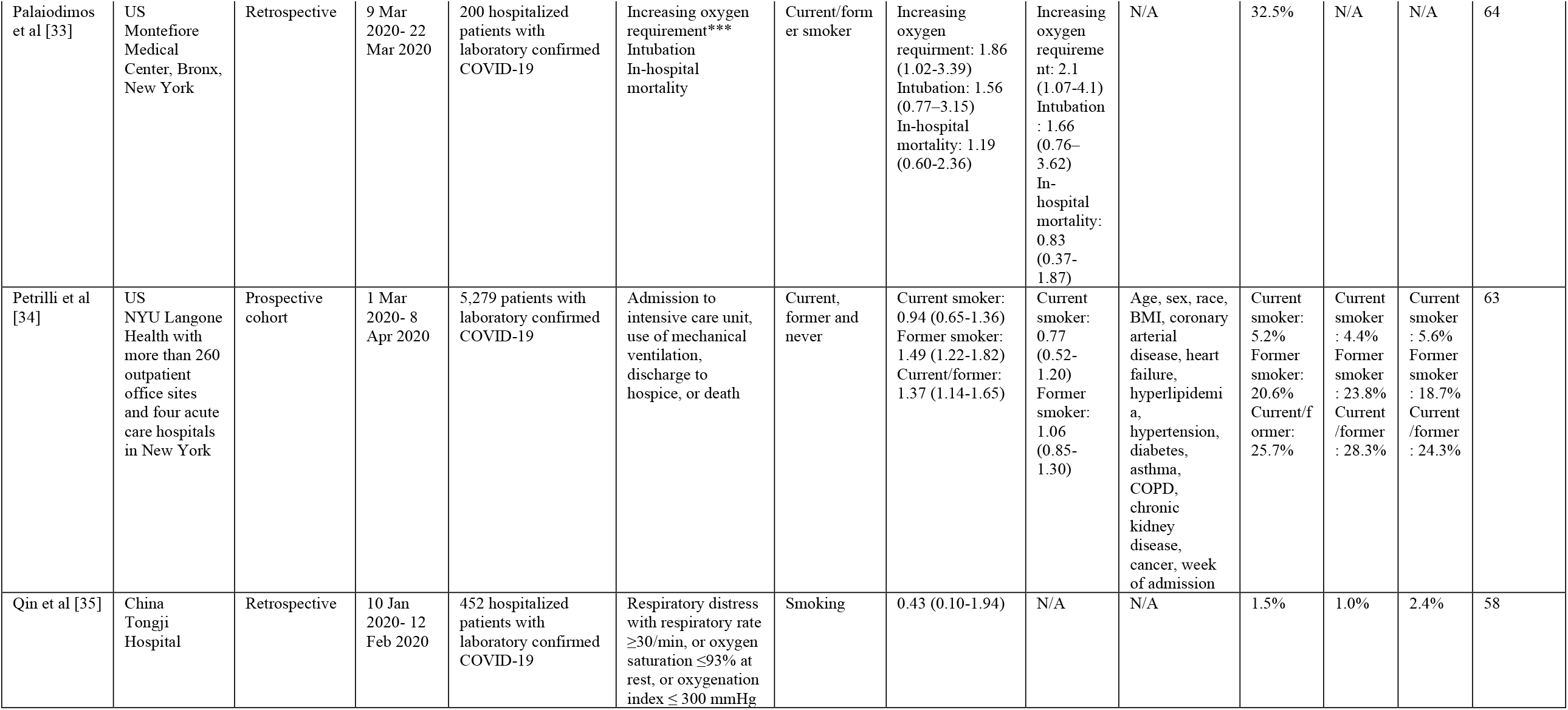
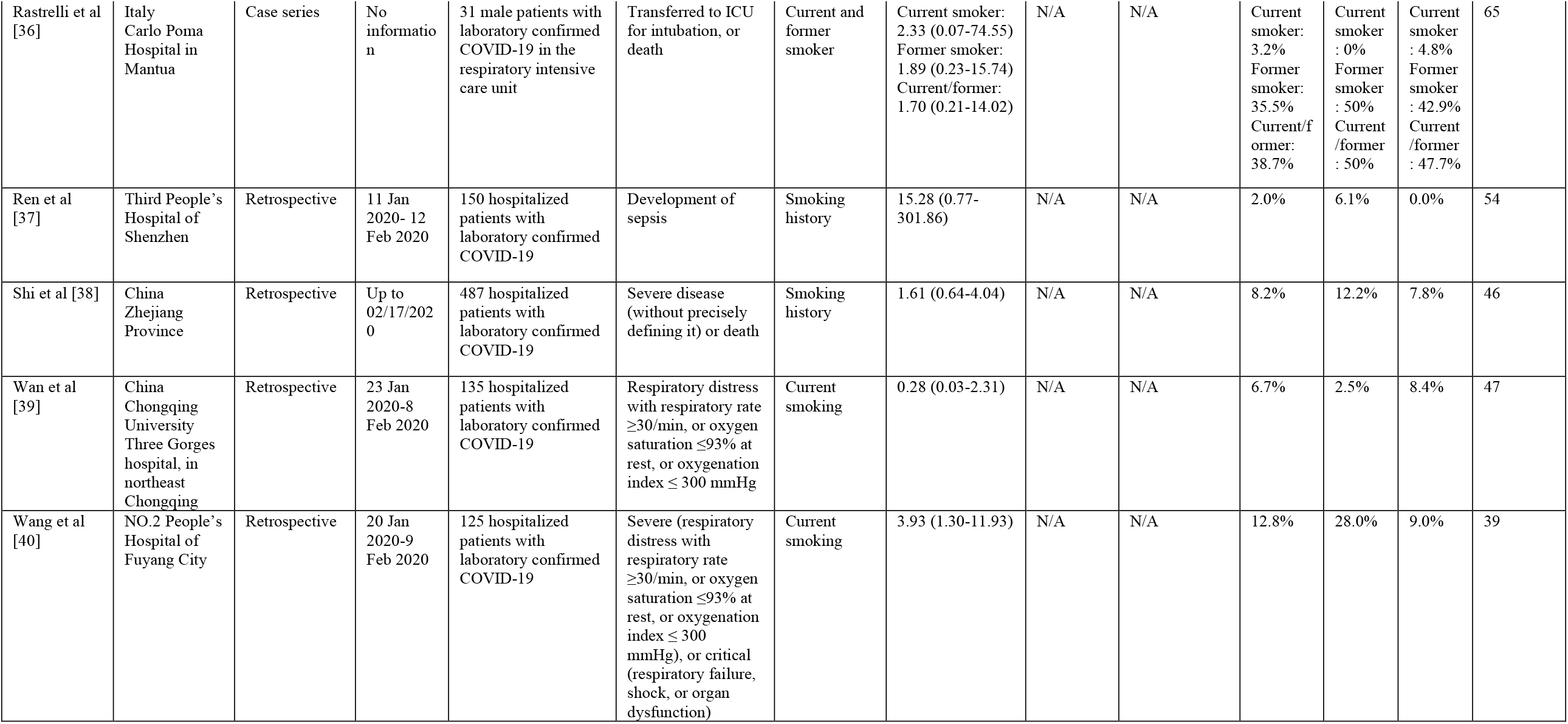
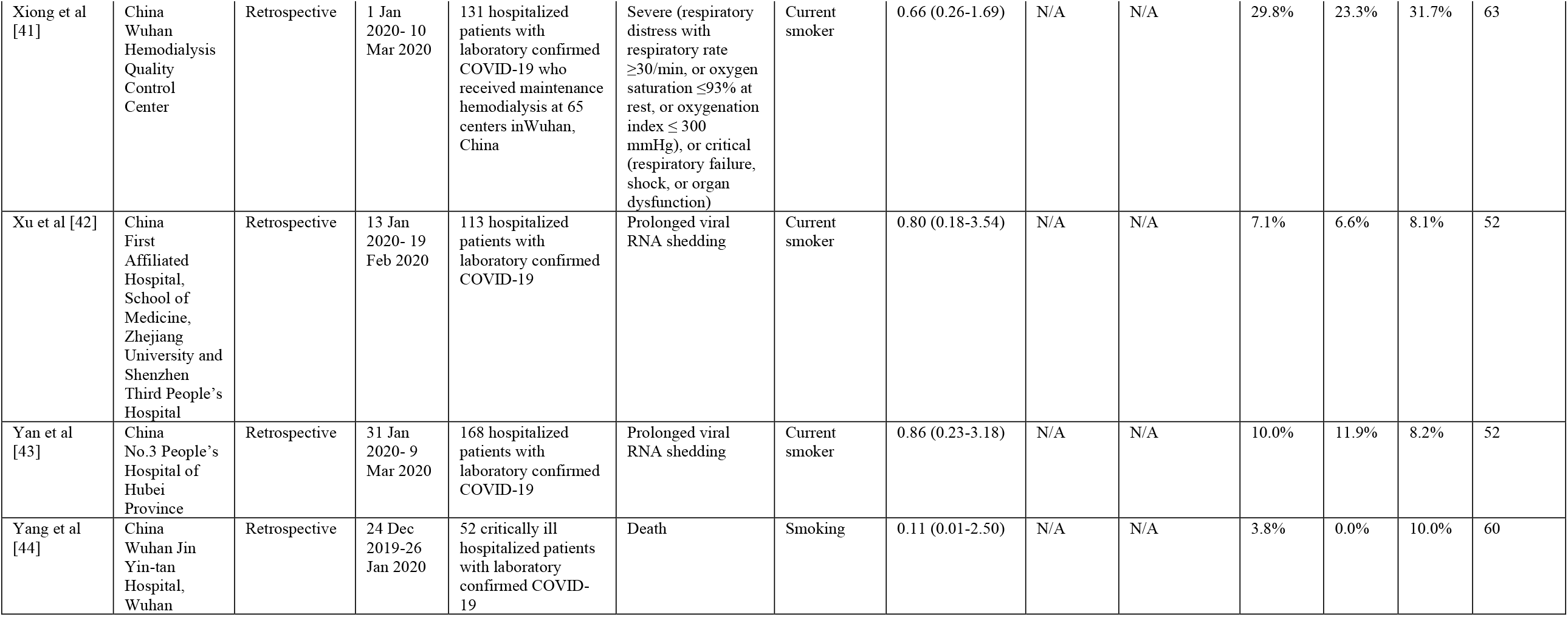
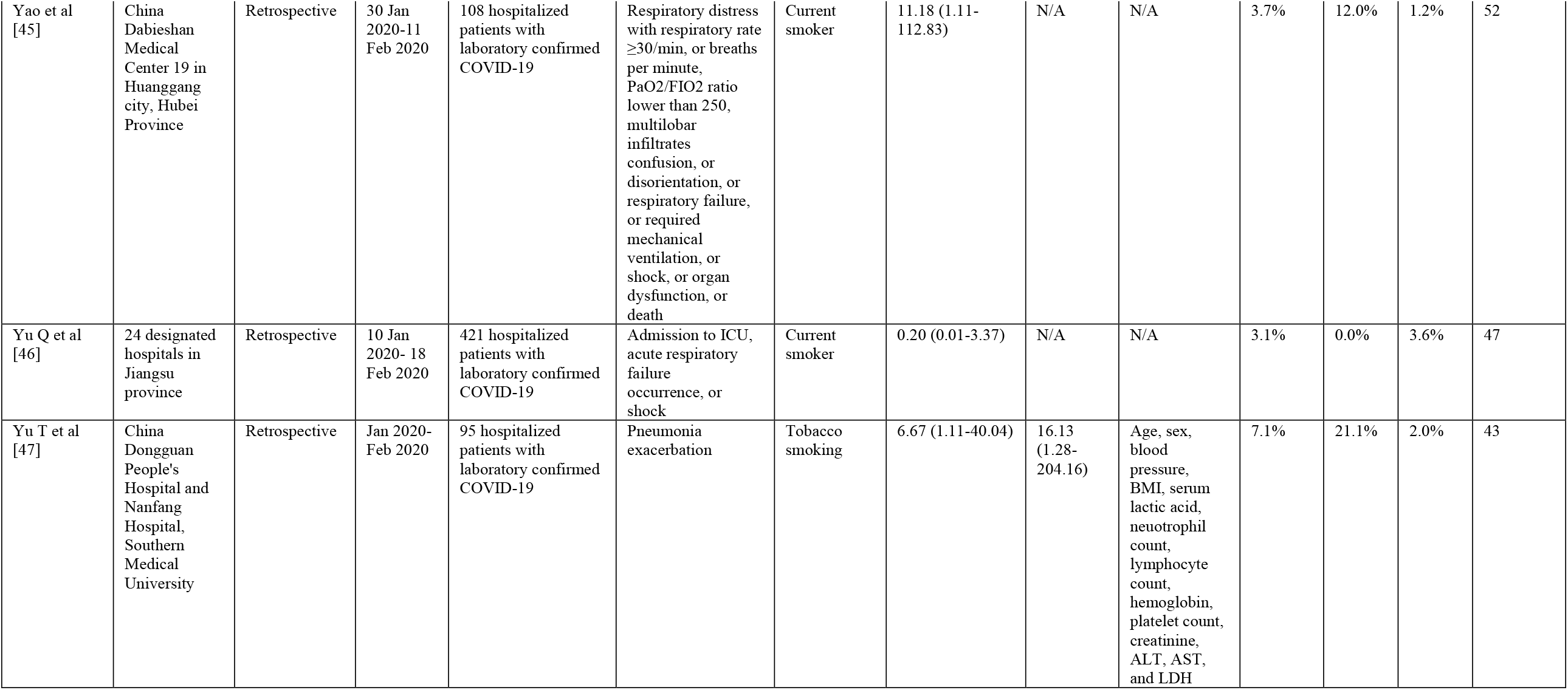
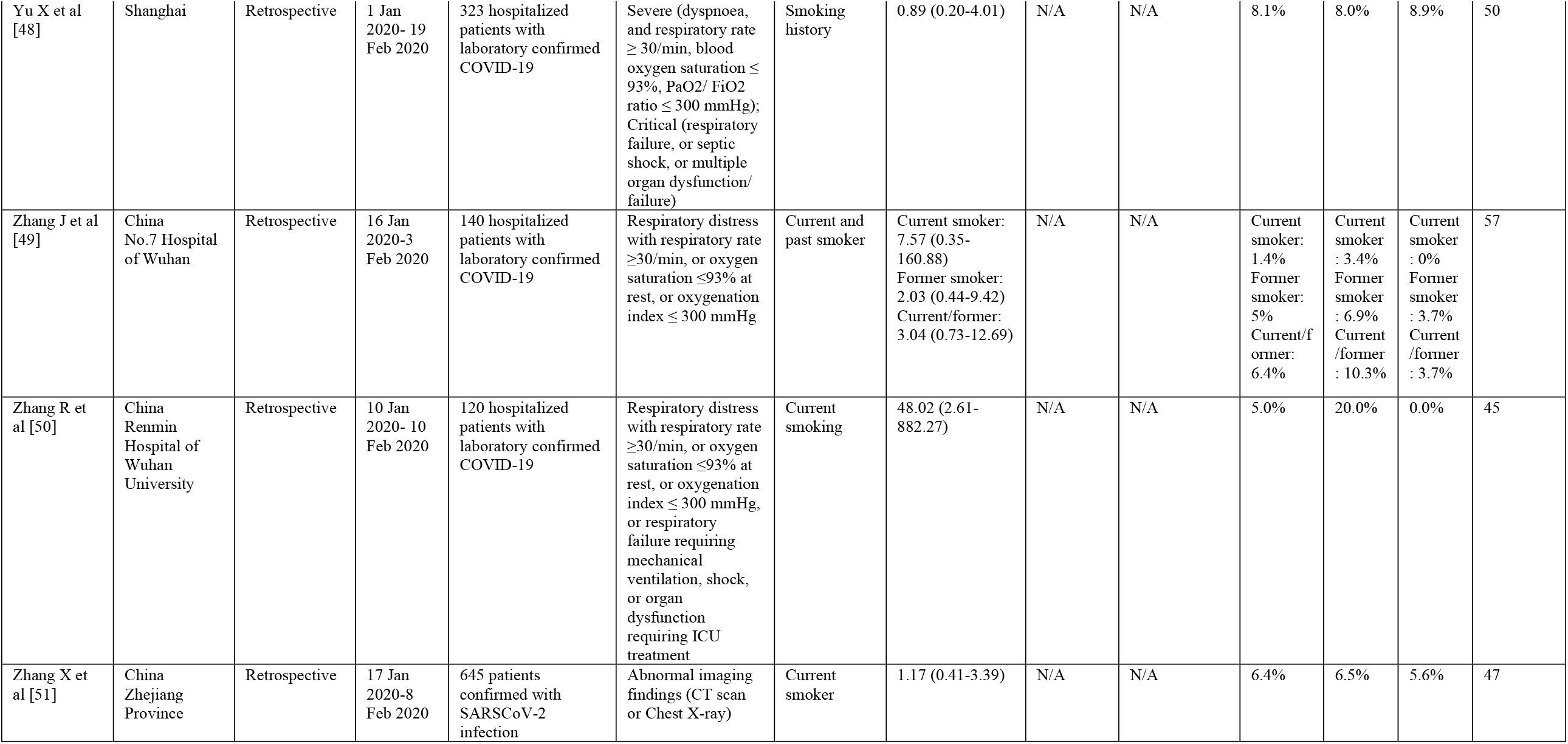
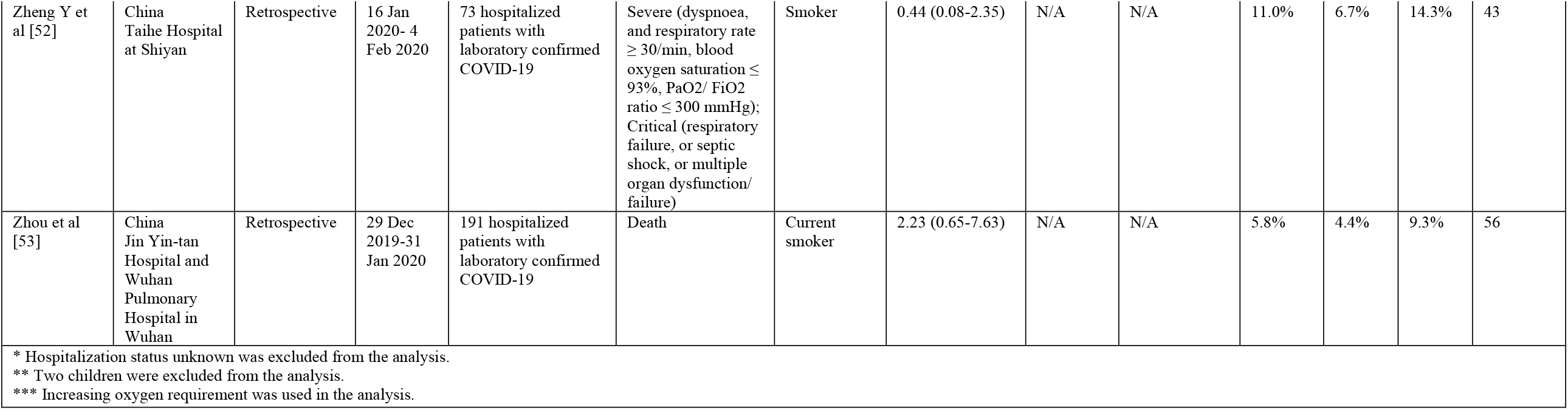
Summary of Studies

| **Table A1 Summary of Studies** |  |  |  |  |  |  |  |  |  |  |  |  |  |
| --- | --- | --- | --- | --- | --- | --- | --- | --- | --- | --- | --- | --- | --- |
| Author | Setting | Study type | Time of data collection | Population | Definition of disease progression | Smoking status | Odds of disease progression (95%CI) |  | Adjustment | Smoking prevalence |  |  | Patients' mean age |
|  |  |  |  |  |  |  | Unadjusted | Adjusted |  | Overall | More severe group | Less severe group |  |
| Beyrouti et al [7] | UK National Hospital for Neurology and Neurosurgery, Queen Square, London | Case series | 1 Apr 2020-16 Apr 2020 | 6 confirmed COVID-19 patients with acute ischemic stroke | ICU admission | Smoking | 0.47 (0.01, 16.89) | N/A | N/A | 16.7% | N/A | 25.0% | 70 |
| Buckner et al[8] | US University of Washington affiliated hospitals | Retrospective | 2 Mar 2020-26 Mar 2020 | 105 hospitalized patients with laboratory confirmed COVID-19 | A composite endpoint of admission to an intensive care unit (ICU) or death | Smoking (ever) | 0.89 (0.34, 2.35) | N/A | N/A | 25.6% | 24.4% | 26.7% | 69 |
| CDC COVID-19 Response Team [9] | US | Retrospective | 12 Feb 2020-28 Mar 2020 | 7,162 confirmed COVID-19 patients with completed information* | ICU admission | Current and former smoker | Current smoker: 0.86 (0.35-2.13)<br>Former smoker: 3.76 (2.53-5.59)<br>Current/former: 2.60 (1.82-3.73) | N/A | N/A | Current smoker: 1.3%<br>Former smoker: 2.3%<br>Current/former: 3.6% | Current smoker : 0.5%<br>Former smoker : 3.1%<br>Current /former : 3.6% | Current smoker : 1.3%<br>Former smoker : 2.0%<br>Current /former : 3.4% | N/A |
| Chen Q et al [10] | China Taizhou Public Health Medical Center, Zhejiang | Retrospective | 1 Jan 2020- 11 Mar 2020 | 145 hospitalized patients with laboratory confirmed COVID-19 | Respiratory distress with respiratory rate $\geq 30$/min, or oxygen saturation $\leq 93\%$ at rest, or oxygenation index $\leq 300$ mmHg | Smoking history | 0.56 (0.15-2.10) | N/A | N/A | 10.3% | 7.0% | 11.8% | 48 |
| Chen T et al [11] | China Tongji Hospital | Retrospective | 13 Jan 2020-13 Feb 2020 | 799 moderately to severely ill or critically ill patients with confirmed covid-19 | Death | Current and former smoker | Current smoker: 2.03 (0.63-6.58)<br>Former smoker: 0.58 (0.11-3.05)<br>Current/former: 1.31 (0.51-3.33) | N/A | N/A | Current smoker: 4.4%<br>Former smoker: 2.6%<br>Current/f ormer: 6.9% | Current smoker : 6.2%<br>Former smoker : 1.8%<br>Current /former : 8.0% | Current smoker : 3.1%<br>Former smoker : 3.1%<br>Current /former : 6.2% | 62 |
| **Author** | **Setting** | **Study type** | **Time of data collection** | **Population** | **Definition of disease progression** | **Smoking status** | **Odds of disease progression (95%CI)** |  | **Adjustment** | **Smoking prevalence** |  |  | **Patients' mean age** |
|  |  |  |  |  |  |  | **Unadjusted** | **Adjusted** |  | **Overall** | **More severe group** | **Less severe group** |  |
| Dong et al [12] | China Zhongnan Hospital of Wuhan University, No. 7 hospital of Wuhan, and Wuhan Children's Hospital | Case series | 20 Jan 2020-3 Mar 2020 | 9 adults hospitalized patients with laboratory confirmed COVID-19** | Bilateral pneumonia, respiratory distress, required mechanical ventilation, ICU care, or hospitalized > 10 days. | Current smoker | 1.15 (0.03-38.88) | N/A | N/A | 11.1% | 14.3% | 0.0% | 44 |
| Feng et al [13] | Jinyintan Hospital in Wuhan, Shanghai Public Health Clinical Center in Shanghai and Tongling People's Hospital in Anhui Province | Retrospective | 1 Jan 2020-15 Feb 2020 | 476 hospitalized patients with laboratory confirmed COVID-19 | Severe (respiratory distress with respiratory rate $\geq 30$/min, or oxygen saturation $\leq 93\%$ at rest, or oxygenation index $\leq 300$ mmHg), or critical (respiratory failure, shock, or organ dysfunction) | Smoking | 1.91 (1.00-3.65) | N/A | N/A | 9.2% | 13.7% | 7.7% | 53 |
| Gayam et al [14] | Interfaith Medical Center, Brooklyn, New York | Case series | 1 Mar 2020-15 April 2020 | 6 confirmed hospitalized COVID-19 patients coinfectd with Mycoplasma pneumoniae | Required intensive mechanical ventilator | Current smoker | 0.47 (0.01-16.89) | N/A | N/A | 33.3% | 0.0% | 40.0% | 57 |
| Giacomelliet al [15] | Infectious Diseases and Intensive Care units of Luigi Sacco Hospital, Milan | Prospective cohort | 21 Feb 2020-19 Mar 2020 | 233 hospitalized patients with laboratory confirmed COVID-19 | Death | History of smoking | 1.37 (0.70-2.68) | N/A | N/A | 30.0% | 35.4% | 28.6% | 61 |
| **Author** | **Setting** | **Study type** | **Time of data collection** | **Population** | **Definition of disease progression** | **Smoking status** | **Odds of disease progression (95%CI)** |  | **Adjustment** | **Smoking prevalence** |  |  | **Patients' mean age** |
|  |  |  |  |  |  |  | **Unadjusted** | **Adjusted** |  | **Overall** | **More severe group** | **Less severe group** |  |
| Goyal et al [16] | US 862-bed quaternary referral center and an affiliated 180-bed nonteaching community hospital in Manhattan, New York | Retrospective | 3 Mar 2020-27 Mar 2020 | 393 hospitalized patients with COVID-19 diagnosis | Required intensive mechanical ventilator | Current smoker | 0.86 (0.32-2.29) | N/A | N/A | 5.1% | 4.6% | 5.3% | 62 |
| Guan et al [17] | 552 hospitals in 30 provinces, autonomous regions, and municipalities | Retrospective | 11 Dec 2019-29 Jan 2020 | 1,099 patients (both hospitalized and outpatient) with laboratory confirmed COVID-19 | ICU admission, the use of mechanical ventilation, or death | Current, former and never | Current smoker: 2.84 (1.57-5.14)<br>Former smoker: 6.27 (2.20-17.90)<br>Current/former: 3.25 (1.89-5.59) | N/A | N/A | Current smoker: 12.6%<br>Former smoker: 1.9%<br>Current/former: 14.6% | Current smoker : 25.8%<br>Former smoker : 7.6%<br>Current /former : 33.3% | Current smoker : 11.8%<br>Former smoker : 1.6%<br>Current /former : 13.3% | 47 |
| Guo et al [18] | Seventh Hospital of Wuhan City | Retrospective | 23 Jan 2020-23 Feb 2020 | 187 hospitalized patients with laboratory confirmed COVID-19 | Acute myocardial injury | Smoking | 1.75 (0.64-4.80) | N/A | N/A | 9.6% | 13.5% | 8.1% | 59 |
| **Author** | **Setting** | **Study type** | **Time of data collection** | **Population** | **Definition of disease progression** | **Smoking status** | **Odds of disease progression (95%CI)** |  | **Adjustment** | **Smoking prevalence** |  |  | **Patients' mean age** |
|  |  |  |  |  |  |  | **Unadjusted** | **Adjusted** |  | **Overall** | **More severe group** | **Less severe group** |  |
| Hu et al [19] | China<br>Tianyou Hospital in<br>Wuhan | Retrospective | 8 Jan<br>2020- 20<br>Feb 2020 | 323 hospitalized<br>patients with<br>COVID-19 diagnosis | Death, progression from non-severe to severe (patients showed features of nonsevere patients plus respiratory distress with RR $\geq 30$ breaths/min, oxygen saturation $\leq 93\%$, arterial partial pressure of oxygen/oxygen concentration $\leq 300$ mmHg) or critical (patients showed respiratory failure requiring ventilatory support, as well as shock and organ dysfunction requiring intensive care) or severe to critical status, and/or maintenance of severe or critical status | Smoking history | 2.12 (1.00-4.47) | 3.46 (1.18-10.17) | Age, hypnotics, diagnosis at critical status, diabetes, hypertensive troponin I, white blood cell count, neutrophil count | 11.8% | 15.1% | 7.9% | 61 |
| Huang C et al [20] | China<br>Jin Yin-tan Hospital in<br>Wuhan | Retrospective | 16 Dec<br>2019-2<br>Jan 2020 | 41 hospitalized<br>patients with<br>laboratory confirmed<br>COVID-19 | ICU admission | Current smoking | 0.27 (0.01-5.62) | N/A | N/A | 7.3% | 0.0% | 10.7% | 49 |
| Huang R et al [21] | China<br>8 designated hospitals in 8<br>cities of<br>Jiangsu Province | Retrospective | 22 Jan<br>2020- 10<br>Feb 2020 | 202 hospitalized<br>patients with<br>laboratory confirmed<br>COVID-19 | Severe (respiratory distress with respiratory rate $\geq 30$/min, or oxygen saturation $\leq 93\%$ at rest, or oxygenation index $\leq 300$ mmHg), or critical (respiratory failure, shock, or organ dysfunction) | Smoking history | 1.12 (0.24, 5.22) | N/A | N/A | 7.9% | 8.7% | 7.8% | 44 |
| **Author** | **Setting** | **Study type** | **Time of data collection** | **Population** | **Definition of disease progression** | **Smoking status** | **Odds of disease progression (95%CI)** |  | **Adjustment** | **Smoking prevalence** |  |  | **Patients' mean age** |
|  |  |  |  |  |  |  | **Unadjusted** | **Adjusted** |  | **Overall** | **More severe group** | **Less severe group** |  |
| Hur et al [22] | US<br>10 hospitals in the Chicago metropolitan area | Retrospective | 1 Mar 2020- 8 April 2020 | 486 hospitalized patients with laboratory confirmed COVID-19 | Intubation and prolonged intubation for acute respiratory failure | Current/former smoker | 1.47 (0.98-2.22) | N/A | N/A | 33.5% | 39.9% | 31.0% | 59 |
| Ji et al [23] | China<br>Fuyang Second People's Hospital and the Fifth Medical Center of Chinese PLA General Hospital | Retrospective | 20 Jan 2020-22 Feb 2020 | 208 hospitalized patients with laboratory confirmed COVID-19 | Progression to respiratory distress with respiratory rate $\geq 30$/min, or oxygen saturation $\leq 93\%$ at rest, or oxygenation index $\leq 300$ mmHg, or requirement of mechanical ventilator | Smoke | 2.10 (0.75-5.93) | N/A | N/A | 9.1% | 15.0% | 7.7% | 44 |
| Kim et al [24] | Korea | Retrospective | 19 Jan 2020-17 Feb 2020 | 28 confirmed COVID-10 cases nationwide | Required oxygen supplement | Smoking | 3.17 (0.39-25.58) | N/A | N/A | 18.5% | 33.3% | 13.6% | 43 |
| Klang et al [25] | US<br>5 hospital campuses of the Mount Sinai hospital, New York | Retrospective | 1 Mar 2020- 17 May 2020 | 3,406 hospitalized patients with laboratory confirmed COVID-19 | Death | Smoking | age $\leq 50$: 1.97 (1.01-3.85)<br>age $> 50$: 1.13 (0.95-1.35) | age $\leq 50$: 1.7 (0.8-1.2)<br>age $> 50$: 1.0 (0.8-1.2) | Age, sex, coronary arterial disease, congestive heart failure, hypertension, diabetes, hyperlipidemia, chronic kidney disease, cancer, intubation, BMI, and race | age $\leq 50$: 13.3%<br>age $> 50$: 25.3% | age $\leq 50$: 21.7%<br>age $> 50$: 26.8% | age $\leq 50$: 12.3%<br>age $> 50$: 24.4% | age $\leq 50$: 41<br>age $> 50$: 71 |
| Author | Setting | Study type | Time of data collection | Population | Definition of disease progression | Smoking status | Odds of disease progression (95%CI) |  | Adjustment | Smoking prevalence |  |  | Patients' mean age |
|  |  |  |  |  |  |  | Unadjusted | Adjusted |  | Overall | More severe group | Less severe group |  |
| Li X et al[26] | China Tongji Hospital | Retrospective | 26 Jan 2020-5 Feb 2020 | 548 hospitalized patients with laboratory confirmed COVID-19 | Respiratory distress with respiratory rate $\geq 30$/min, or breaths per minute, PaO<sub>2</sub>/FIO<sub>2</sub> ratio lower than 250, multilobar infiltrates confusion, or disorientation, or respiratory failure, or required mechanical ventilation, or septic shock | Current, former and never | Current smoker: 0.87 (0.46-1.66)<br>Former smoker: 2.04 (1.12-3.73)<br>Current/former: 1.38 (0.88-2.17) | N/A | N/A | Current smoker: 7.5%<br>Former smoker: 9.4%<br>Current/former: 16.9% | Current smoker : 6.8%<br>Former smoker : 12.5%<br>Current /former : 19.2% | Current smoker : 8.2%<br>Former smoker : 6.5%<br>Current /former : 14.7% | 60 |
| Li YK et al[27] | China Department of Thoracic Surgery of Tongji Hospital affiliated with Tongji Medical College of Huazhong University of Science and Technology | Retrospective | 1 Jan 2020- 20 Feb 2020 | 25 hospitalized patients with laboratory confirmed COVID-19 | Severe (respiratory distress with respiratory rate $\geq 30$/min, or oxygen saturation $\leq 93\%$ at rest, or oxygenation index $\leq 300$ mmHg), or critical (respiratory failure, shock, or organ dysfunction) | Smoking | 8.75 (1.21-63.43) | N/A | N/A | 28.0% | 55.6% | 12.5% | 51 |
| Liang et al[28] | China 575 hospitals in 31 provincial administrative regions | Retrospective | 21 Nov 2019- 31 Jan 2020 | 1,590 hospitalized patients with laboratory confirmed COVID-19 | Admission to the intensive care unit (ICU), invasive ventilation, or death | Current/former and never | 2.34 (1.36-4.02) | N/A | N/A | 7.0% | 13.7% | 6.4% | 49 |
| **Author** | **Setting** | **Study type** | **Time of data collection** | **Population** | **Definition of disease progression** | **Smoking status** | **Odds of disease progression (95%CI)** |  | **Adjustment** | **Smoking prevalence** |  |  | **Patients' mean age** |
|  |  |  |  |  |  |  | **Unadjusted** | **Adjusted** |  | **Overall** | **More severe group** | **Less severe group** |  |
| Liu et al [29] | China<br>Three tertiary hospitals in Wuhan | Retrospective | 30 Dec 2019-15 Jan 2020 | 78 hospitalized patients with laboratory confirmed COVID-19 | Respiratory distress, respiratory failure, required mechanical ventilation and ICU care, or death | History of smoking | 12.19 (1.76-84.31) | 14.29 (1.58-25.0) | Age, maximum temperature at admission, respiratory failure, severe illness, albumin, creatinine, procalcitonin, and C-reactive protein level | 6.4% | 27.3% | 3.0% | 38 |
| Mehra et al [30] | 11 Countries in Asia, Europe, and North America (China, Japan, South Korea, Turkey, Spain, Italy, Germany, France, UK, Canada, and US) | Retrospective | 20 Dec 2020- 15 Mar 2020 | 8,910 hospitalized patients with laboratory confirmed COVID-19 in 169 hospitals | Death | Current and former smoker | Current smoker: 1.75 (1.27-2.41)<br>Former smoker: 1.00 (0.78-1.27)<br>Current/former: 1.18 (0.96-1.45) | Current smoker: 1.79 (1.29-2.47) | Age, sex, coronary arterial disease, congestive heart failure, arrhythmia, COPD, receiving ACE inhibitor, receiving ARB, receiving statin | Current smoker: 5.5%<br>Former smoker: 16.8%<br>Current/former: 22.3% | Current smoker : 5.3%<br>Former smoker : 16.8%<br>Current /former : 22.1% | Current smoker : 8.9%<br>Former smoker : 16.1%<br>Current /former : 25.0% | 49 |
| Mo et al [31] | China<br>Zhongnan hospital of Wuhan University | Retrospective | 1 Jan 2020-5 Feb 2020 | 155 hospitalized patients with laboratory confirmed COVID-19 | Did not improve after treatment, status changed to severe, or hospitalized > 10 days | Current smoking | 1.68 (0.30-9.45) | N/A | N/A | 3.9% | 4.7% | 2.9% | 54 |
| Morassi et al [32] | Italy<br>Fondazione Poliambulanza Hospital, Brescia, Italy and Udine University Hospital, Udine | Case series | 16 Mar 2020- 5 Apr 2020 | 6 hospitalized patients with laboratory confirmed COVID-19 who developed clinical and neuroimaging evidence of acute stroke | Death | Smoker | 1.00 (0.03-40.28) | N/A | N/A | 16.7% | 20.0% | 0.0% | 69 |

| Table A1 Summary of Studies |  |  |  |  |  |  |  |  |  |  |  |  |  |
| --- | --- | --- | --- | --- | --- | --- | --- | --- | --- | --- | --- | --- | --- |
| Author | Setting | Study type | Time of data collection | Population | Definition of disease progression | Smoking status | Odds of disease progression (95%CI) |  | Adjustment | Smoking prevalence |  |  | Patients' mean age |
|  |  |  |  |  |  |  | Unadjusted | Adjusted |  | Overall | More severe group | Less severe group |  |
| Palaiodimos et al [33] | US Montefiore Medical Center, Bronx, New York | Retrospective | 9 Mar 2020- 22 Mar 2020 | 200 hospitalized patients with laboratory confirmed COVID-19 | Increasing oxygen requirement***<br>Intubation<br>In-hospital mortality | Current/former smoker | Increasing oxygen requirement: 1.86 (1.02-3.39)<br>Intubation: 1.56 (0.77-3.15)<br>In-hospital mortality: 1.19 (0.60-2.36) | Increasing oxygen requirement: 2.1 (1.07-4.1)<br>Intubation: 1.66 (0.76-3.62)<br>In-hospital mortality: 0.83 (0.37-1.87) | N/A | 32.5% | N/A | N/A | 64 |
| Petrilli et al [34] | US NYU Langone Health with more than 260 outpatient office sites and four acute care hospitals in New York | Prospective cohort | 1 Mar 2020- 8 Apr 2020 | 5,279 patients with laboratory confirmed COVID-19 | Admission to intensive care unit, use of mechanical ventilation, discharge to hospice, or death | Current, former and never | Current smoker: 0.94 (0.65-1.36)<br>Former smoker: 1.49 (1.22-1.82)<br>Current/former: 1.37 (1.14-1.65) | Current smoker: 0.77 (0.52-1.20)<br>Former smoker: 1.06 (0.85-1.30) | Age, sex, race, BMI, coronary arterial disease, heart failure, hyperlipidemia, hypertension, diabetes, asthma, COPD, chronic kidney disease, cancer, week of admission | Current smoker: 5.2%<br>Former smoker: 20.6%<br>Current/former: 25.7% | Current smoker: 4.4%<br>Former smoker: 23.8%<br>Current/former: 28.3% | Current smoker: 5.6%<br>Former smoker: 18.7%<br>Current/former: 24.3% | 63 |
| Qin et al [35] | China Tongji Hospital | Retrospective | 10 Jan 2020- 12 Feb 2020 | 452 hospitalized patients with laboratory confirmed COVID-19 | Respiratory distress with respiratory rate $\geq 30$/min, or oxygen saturation $\leq 93\%$ at rest, or oxygenation index $\leq 300$ mmHg | Smoking | 0.43 (0.10-1.94) | N/A | N/A | 1.5% | 1.0% | 2.4% | 58 |
| Author | Setting | Study type | Time of data collection | Population | Definition of disease progression | Smoking status | Odds of disease progression (95%CI) |  | Adjustment | Smoking prevalence |  |  | Patients' mean age |
|  |  |  |  |  |  |  | Unadjusted | Adjusted |  | Overall | More severe group | Less severe group |  |
| Rastrelli et al [36] | Italy Carlo Poma Hospital in Mantua | Case series | No information | 31 male patients with laboratory confirmed COVID-19 in the respiratory intensive care unit | Transferred to ICU for intubation, or death | Current and former smoker | Current smoker: 2.33 (0.07-74.55)<br>Former smoker: 1.89 (0.23-15.74)<br>Current/former: 1.70 (0.21-14.02) | N/A | N/A | Current smoker: 3.2%<br>Former smoker: 35.5%<br>Current/former: 38.7% | Current smoker : 0%<br>Former smoker : 50%<br>Current /former : 50% | Current smoker : 4.8%<br>Former smoker : 42.9%<br>Current /former : 47.7% | 65 |
| Ren et al [37] | Third People's Hospital of Shenzhen | Retrospective | 11 Jan 2020- 12 Feb 2020 | 150 hospitalized patients with laboratory confirmed COVID-19 | Development of sepsis | Smoking history | 15.28 (0.77-301.86) | N/A | N/A | 2.0% | 6.1% | 0.0% | 54 |
| Shi et al [38] | China Zhejiang Province | Retrospective | Up to 02/17/2020 | 487 hospitalized patients with laboratory confirmed COVID-19 | Severe disease (without precisely defining it) or death | Smoking history | 1.61 (0.64-4.04) | N/A | N/A | 8.2% | 12.2% | 7.8% | 46 |
| Wan et al [39] | China Chongqing University Three Gorges hospital, in northeast Chongqing | Retrospective | 23 Jan 2020-8 Feb 2020 | 135 hospitalized patients with laboratory confirmed COVID-19 | Respiratory distress with respiratory rate $\geq 30$/min, or oxygen saturation $\leq 93\%$ at rest, or oxygenation index $\leq 300$ mmHg | Current smoking | 0.28 (0.03-2.31) | N/A | N/A | 6.7% | 2.5% | 8.4% | 47 |
| Wang et al [40] | NO.2 People's Hospital of Fuyang City | Retrospective | 20 Jan 2020-9 Feb 2020 | 125 hospitalized patients with laboratory confirmed COVID-19 | Severe (respiratory distress with respiratory rate $\geq 30$/min, or oxygen saturation $\leq 93\%$ at rest, or oxygenation index $\leq 300$ mmHg), or critical (respiratory failure, shock, or organ dysfunction) | Current smoking | 3.93 (1.30-11.93) | N/A | N/A | 12.8% | 28.0% | 9.0% | 39 |
| **Author** | **Setting** | **Study type** | **Time of data collection** | **Population** | **Definition of disease progression** | **Smoking status** | **Odds of disease progression (95%CI)** |  | **Adjustment** | **Smoking prevalence** |  |  | **Patients' mean age** |
|  |  |  |  |  |  |  | **Unadjusted** | **Adjusted** |  | **Overall** | **More severe group** | **Less severe group** |  |
| Xiong et al [41] | China<br>Wuhan<br>Hemodialysis<br>Quality<br>Control<br>Center | Retrospective | 1 Jan<br>2020- 10<br>Mar 2020 | 131 hospitalized patients with laboratory confirmed COVID-19 who received maintenance hemodialysis at 65 centers in Wuhan, China | Severe (respiratory distress with respiratory rate $\geq 30$/min, or oxygen saturation $\leq 93\%$ at rest, or oxygenation index $\leq 300$ mmHg), or critical (respiratory failure, shock, or organ dysfunction) | Current smoker | 0.66 (0.26-1.69) | N/A | N/A | 29.8% | 23.3% | 31.7% | 63 |
| Xu et al [42] | China<br>First<br>Affiliated<br>Hospital,<br>School of<br>Medicine,<br>Zhejiang<br>University and<br>Shenzhen<br>Third People's<br>Hospital | Retrospective | 13 Jan<br>2020- 19<br>Feb 2020 | 113 hospitalized patients with laboratory confirmed COVID-19 | Prolonged viral RNA shedding | Current smoker | 0.80 (0.18-3.54) | N/A | N/A | 7.1% | 6.6% | 8.1% | 52 |
| Yan et al [43] | China<br>No.3 People's<br>Hospital of<br>Hubei<br>Province | Retrospective | 31 Jan<br>2020- 9<br>Mar 2020 | 168 hospitalized patients with laboratory confirmed COVID-19 | Prolonged viral RNA shedding | Current smoker | 0.86 (0.23-3.18) | N/A | N/A | 10.0% | 11.9% | 8.2% | 52 |
| Yang et al [44] | China<br>Wuhan Jin<br>Yin-tan<br>Hospital,<br>Wuhan | Retrospective | 24 Dec<br>2019-26<br>Jan 2020 | 52 critically ill hospitalized patients with laboratory confirmed COVID-19 | Death | Smoking | 0.11 (0.01-2.50) | N/A | N/A | 3.8% | 0.0% | 10.0% | 60 |
| **Author** | **Setting** | **Study type** | **Time of data collection** | **Population** | **Definition of disease progression** | **Smoking status** | **Odds of disease progression (95%CI)** |  | **Adjustment** | **Smoking prevalence** |  |  | **Patients' mean age** |
|  |  |  |  |  |  |  | **Unadjusted** | **Adjusted** |  | **Overall** | **More severe group** | **Less severe group** |  |
| Yao et al [45] | China Dabieshan Medical Center 19 in Huanggang city, Hubei Province | Retrospective | 30 Jan 2020-11 Feb 2020 | 108 hospitalized patients with laboratory confirmed COVID-19 | Respiratory distress with respiratory rate $\geq 30$/min, or breaths per minute, PaO<sub>2</sub>/FIO<sub>2</sub> ratio lower than 250, multilobar infiltrates confusion, or disorientation, or respiratory failure, or required mechanical ventilation, or shock, or organ dysfunction, or death | Current smoker | 11.18 (1.11-112.83) | N/A | N/A | 3.7% | 12.0% | 1.2% | 52 |
| Yu Q et al [46] | 24 designated hospitals in Jiangsu province | Retrospective | 10 Jan 2020- 18 Feb 2020 | 421 hospitalized patients with laboratory confirmed COVID-19 | Admission to ICU, acute respiratory failure occurrence, or shock | Current smoker | 0.20 (0.01-3.37) | N/A | N/A | 3.1% | 0.0% | 3.6% | 47 |
| Yu T et al [47] | China Dongguan People's Hospital and Nanfang Hospital, Southern Medical University | Retrospective | Jan 2020-Feb 2020 | 95 hospitalized patients with laboratory confirmed COVID-19 | Pneumonia exacerbation | Tobacco smoking | 6.67 (1.11-40.04) | 16.13 (1.28-204.16) | Age, sex, blood pressure, BMI, serum lactic acid, neutrophil count, lymphocyte count, hemoglobin, platelet count, creatinine, ALT, AST, and LDH | 7.1% | 21.1% | 2.0% | 43 |
| Author | Setting | Study type | Time of data collection | Population | Definition of disease progression | Smoking status | Odds of disease progression (95%CI) |  | Adjustment | Smoking prevalence |  |  | Patients' mean age |
|  |  |  |  |  |  |  | Unadjusted | Adjusted |  | Overall | More severe group | Less severe group |  |
| Yu X et al [48] | Shanghai | Retrospective | 1 Jan 2020- 19 Feb 2020 | 323 hospitalized patients with laboratory confirmed COVID-19 | Severe (dyspnoea, and respiratory rate $\geq 30/\text{min}$, blood oxygen saturation $\leq 93\%$, PaO<sub>2</sub>/ FiO<sub>2</sub> ratio $\leq 300 \text{ mmHg}$); Critical (respiratory failure, or septic shock, or multiple organ dysfunction/ failure) | Smoking history | 0.89 (0.20-4.01) | N/A | N/A | 8.1% | 8.0% | 8.9% | 50 |
| Zhang J et al [49] | China No.7 Hospital of Wuhan | Retrospective | 16 Jan 2020-3 Feb 2020 | 140 hospitalized patients with laboratory confirmed COVID-19 | Respiratory distress with respiratory rate $\geq 30/\text{min}$, or oxygen saturation $\leq 93\%$ at rest, or oxygenation index $\leq 300 \text{ mmHg}$ | Current and past smoker | Current smoker: 7.57 (0.35-160.88)<br>Former smoker: 2.03 (0.44-9.42)<br>Current/former: 3.04 (0.73-12.69) | N/A | N/A | Current smoker: 1.4%<br>Former smoker: 5%<br>Current/former: 6.4% | Current smoker : 3.4%<br>Former smoker : 6.9%<br>Current /former : 10.3% | Current smoker : 0%<br>Former smoker : 3.7%<br>Current /former : 3.7% | 57 |
| Zhang R et al [50] | China Renmin Hospital of Wuhan University | Retrospective | 10 Jan 2020- 10 Feb 2020 | 120 hospitalized patients with laboratory confirmed COVID-19 | Respiratory distress with respiratory rate $\geq 30/\text{min}$, or oxygen saturation $\leq 93\%$ at rest, or oxygenation index $\leq 300 \text{ mmHg}$, or respiratory failure requiring mechanical ventilation, shock, or organ dysfunction requiring ICU treatment | Current smoking | 48.02 (2.61-882.27) | N/A | N/A | 5.0% | 20.0% | 0.0% | 45 |
| Zhang X et al [51] | China Zhejiang Province | Retrospective | 17 Jan 2020-8 Feb 2020 | 645 patients confirmed with SARSCoV-2 infection | Abnormal imaging findings (CT scan or Chest X-ray) | Current smoker | 1.17 (0.41-3.39) | N/A | N/A | 6.4% | 6.5% | 5.6% | 47 |
| **Author** | **Setting** | **Study type** | **Time of data collection** | **Population** | **Definition of disease progression** | **Smoking status** | **Odds of disease progression (95%CI)** |  | **Adjustment** | **Smoking prevalence** |  |  | **Patients' mean age** |
|  |  |  |  |  |  |  | **Unadjusted** | **Adjusted** |  | **Overall** | **More severe group** | **Less severe group** |  |
| Zheng Y et al [52] | China Taihe Hospital at Shiyan | Retrospective | 16 Jan 2020- 4 Feb 2020 | 73 hospitalized patients with laboratory confirmed COVID-19 | Severe (dyspnoea, and respiratory rate $\geq 30$/min, blood oxygen saturation $\leq 93\%$, PaO<sub>2</sub>/ FiO<sub>2</sub> ratio $\leq 300$ mmHg); Critical (respiratory failure, or septic shock, or multiple organ dysfunction/ failure) | Smoker | 0.44 (0.08-2.35) | N/A | N/A | 11.0% | 6.7% | 14.3% | 43 |
| Zhou et al [53] | China Jin Yin-tan Hospital and Wuhan Pulmonary Hospital in Wuhan | Retrospective | 29 Dec 2019-31 Jan 2020 | 191 hospitalized patients with laboratory confirmed COVID-19 | Death | Current smoker | 2.23 (0.65-7.63) | N/A | N/A | 5.8% | 4.4% | 9.3% | 56 |
| * Hospitalization status unknown was excluded from the analysis.<br>** Two children were excluded from the analysis.<br>*** Increasing oxygen requirement was used in the analysis. |  |  |  |  |  |  |  |  |  |  |  |  |  |

**Table A2.** Risk of Bias of Studies. (2=high risk, 1=intermediate risk, 0=low risk)

|  |  |  |  |  |  |  |
| --- | --- | --- | --- | --- | --- | --- |
| **Study** | **Study population** | **Exposure measurement** | **Outcome assessment** | **Measurement of confounders** | **Adequate follow-up** | **Average score** |
| Beyrouti et al [7] | 2 | 2 | 0 | 2 | 0 | 1.2 |
| Buckner et al [8] | 0 | 2 | 0 | 2 | 0 | 0.8 |
| CDC COVID-19 [9] | 0 | 0 | 0 | 2 | 0 | 0.4 |
| Chen Q et al [10] | 0 | 2 | 0 | 2 | 0 | 0.8 |
| Chen T et al [11] | 0 | 0 | 0 | 2 | 0 | 0.4 |
| Dong et al [12] | 2 | 2 | 2 | 2 | 0 | 1.6 |
| Feng et al [13] | 0 | 2 | 0 | 2 | 0 | 0.8 |
| Gayam et al [14] | 2 | 2 | 0 | 2 | 0 | 1.2 |
| Giacomelli et al [15] | 0 | 2 | 0 | 2 | 0 | 0.8 |
| Goyal et al [16] | 0 | 2 | 0 | 2 | 0 | 0.8 |
| Guan et al [17] | 0 | 0 | 0 | 2 | 0 | 0.4 |
| Guo et al [18] | 0 | 2 | 0 | 2 | 0 | 0.8 |
| Hu et al [19] | 0 | 2 | 0 | 0 | 0 | 0.4 |
| Huang C et al [20] | 1 | 2 | 0 | 2 | 0 | 1 |
| Huang R et al [21] | 0 | 2 | 0 | 2 | 0 | 0.8 |
| Hur et al [22] | 0 | 1 | 0 | 2 | 0 | 0.6 |
| Ji Dong et al [23] | 0 | 2 | 0 | 2 | 0 | 0.8 |
| Kim et al [24] | 1 | 2 | 0 | 2 | 0 | 1 |
| Klang et al [25] | 0 | 2 | 0 | 0 | 0 | 0.4 |
| Li X et al [26] | 0 | 0 | 0 | 2 | 0 | 0.4 |
| Li YK et al [27] | 1 | 2 | 0 | 2 | 0 | 1 |
| Liang et al [28] | 0 | 1 | 0 | 2 | 0 | 0.6 |
| Liu et al [29] | 0 | 2 | 0 | 0 | 0 | 0.4 |
| Mehra et al [30] | 0 | 0 | 0 | 0 | 0 | 0 |
| Mo et al [31] | 0 | 2 | 0 | 2 | 0 | 0.8 |
| Morassi et al [32] | 2 | 2 | 0 | 2 | 0 | 1.2 |
| Palaiodimos et al [33] | 0 | 1 | 0 | 0 | 0 | 0.2 |
| Petrilli et al [34] | 0 | 0 | 0 | 0 | 0 | 0 |
| Qin et al [35] | 0 | 2 | 0 | 2 | 0 | 0.8 |
| Rastrelli et al [36] | 1 | 0 | 0 | 2 | 2 | 1 |
| Ren et al [37] | 0 | 2 | 0 | 2 | 0 | 0.8 |
| Shi et al [38] | 0 | 2 | 0 | 2 | 2 | 1.2 |
| Wan et al [39] | 0 | 2 | 0 | 2 | 0 | 0.8 |
| Wang et al [40] | 0 | 2 | 0 | 2 | 0 | 0.8 |
| Xiong et al [41] | 0 | 2 | 0 | 2 | 0 | 0.8 |
| Xu et al [42] | 0 | 2 | 0 | 2 | 0 | 0.8 |
| Yan et al [43] | 0 | 2 | 0 | 2 | 0 | 0.8 |
| **Study** | **Study population** | **Exposure measurement** | **Outcome assessment** | **Measurement of confounders** | **Adequate follow-up** | **Average score** |
| Yang et al [44] | 0 | 2 | 0 | 2 | 0 | 0.8 |
| Yao et al [45] | 0 | 2 | 0 | 2 | 0 | 0.8 |
| Yu Q et al [46] | 0 | 2 | 0 | 2 | 0 | 0.8 |
| Yu T et al [47] | 0 | 2 | 0 | 0 | 2 | 0.8 |
| Yu X et al [48] | 0 | 2 | 0 | 2 | 0 | 0.8 |
| Zhang J et al [49] | 0 | 0 | 0 | 2 | 0 | 0.4 |
| Zhang R et al [50] | 0 | 2 | 0 | 2 | 0 | 0.8 |
| Zhang X et al [51] | 0 | 2 | 0 | 2 | 0 | 0.8 |
| Zheng Y et al [52] | 0 | 2 | 0 | 2 | 0 | 0.8 |
| Zhou et al [53] | 0 | 2 | 0 | 2 | 0 | 0.8 |

**Figure A1.**
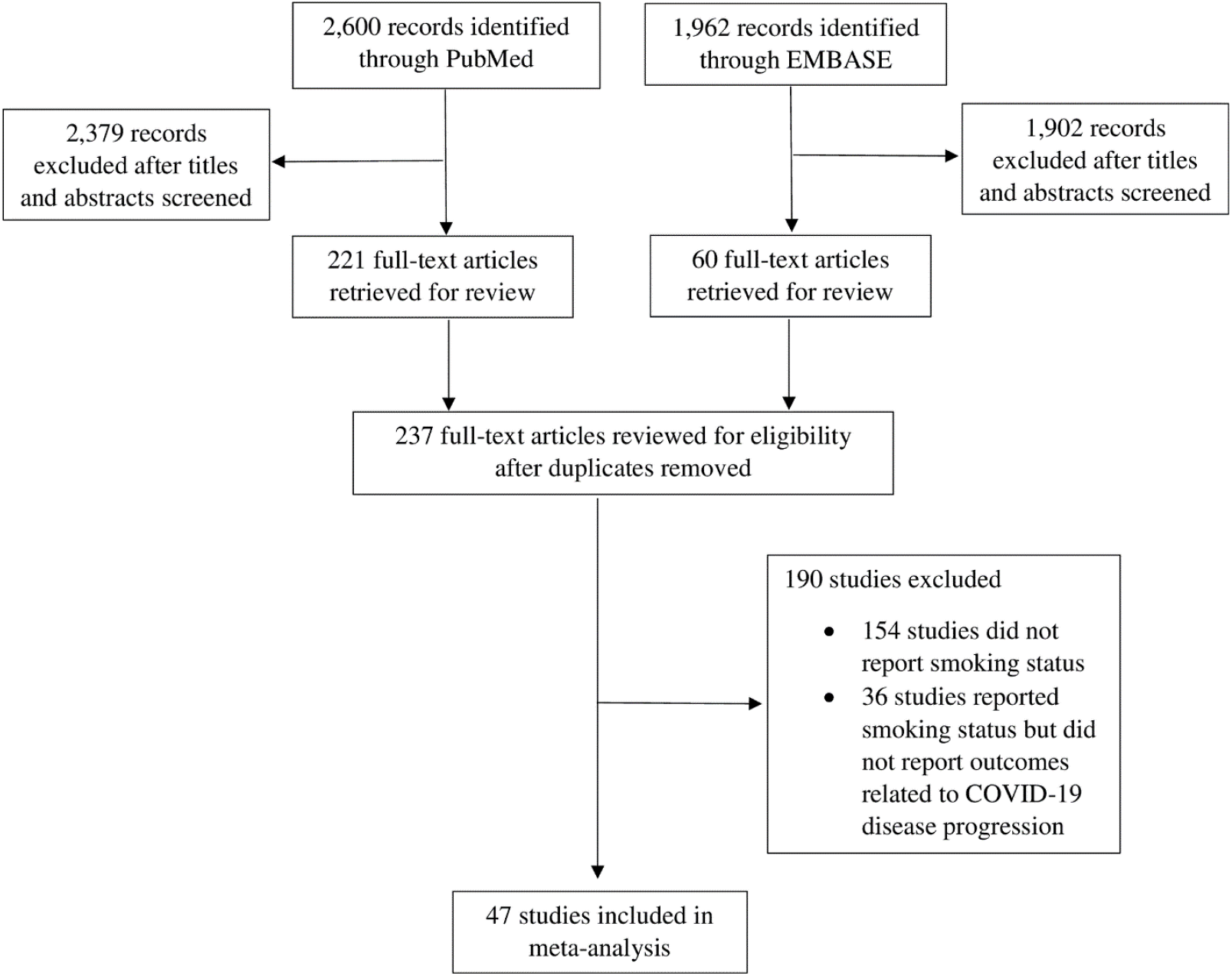
PRISMA diagram

**Figure A2.**
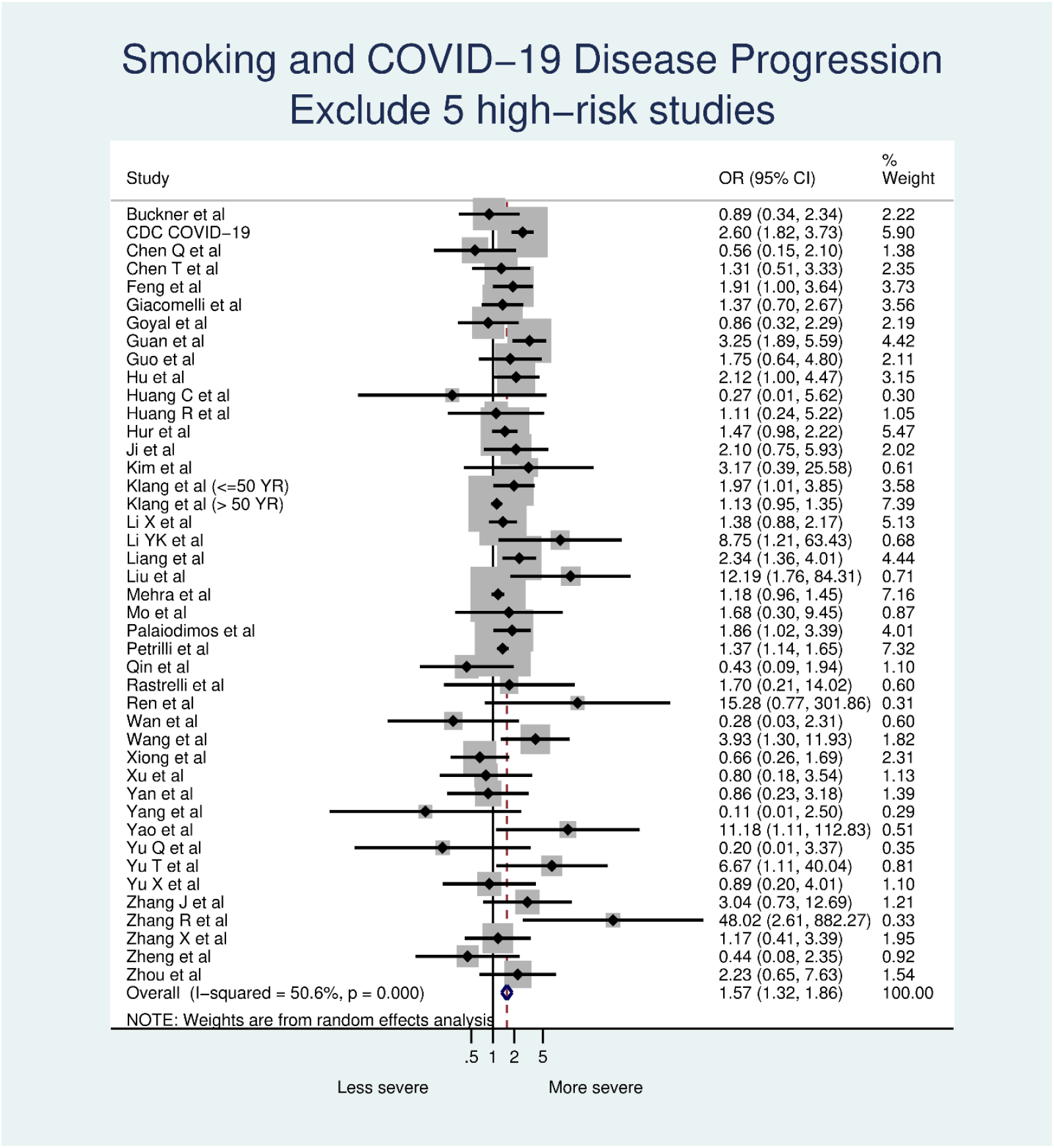
Smoking and COVID-19 disease progression, dropping 5 studies with high risk of bias scores.

**Figure A3.**
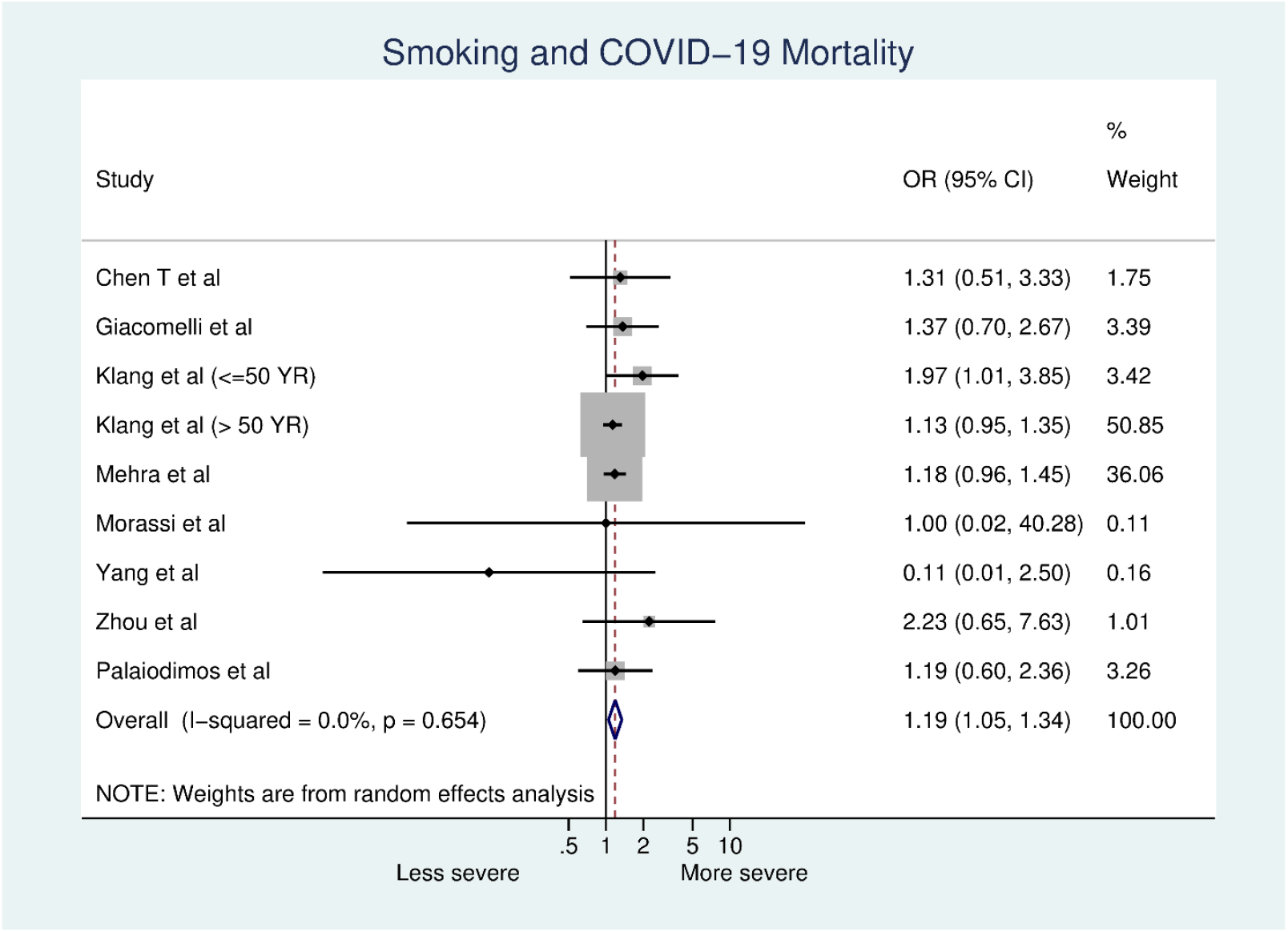
Smoking and COVID-19 mortality

**Figure A4.**
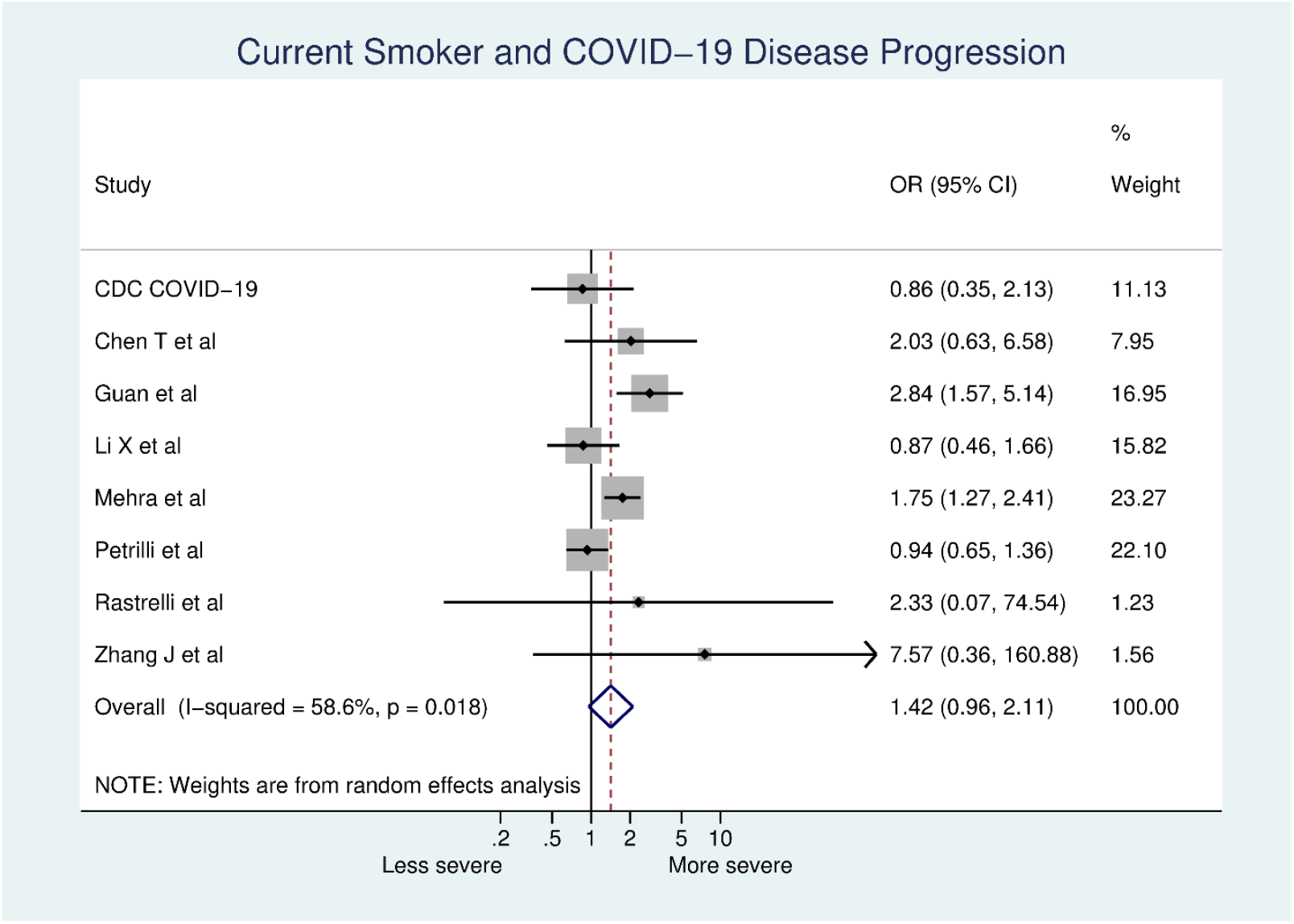
Current smokers vs. never smokers and COVID-19 disease progression.

**Figure A5.**
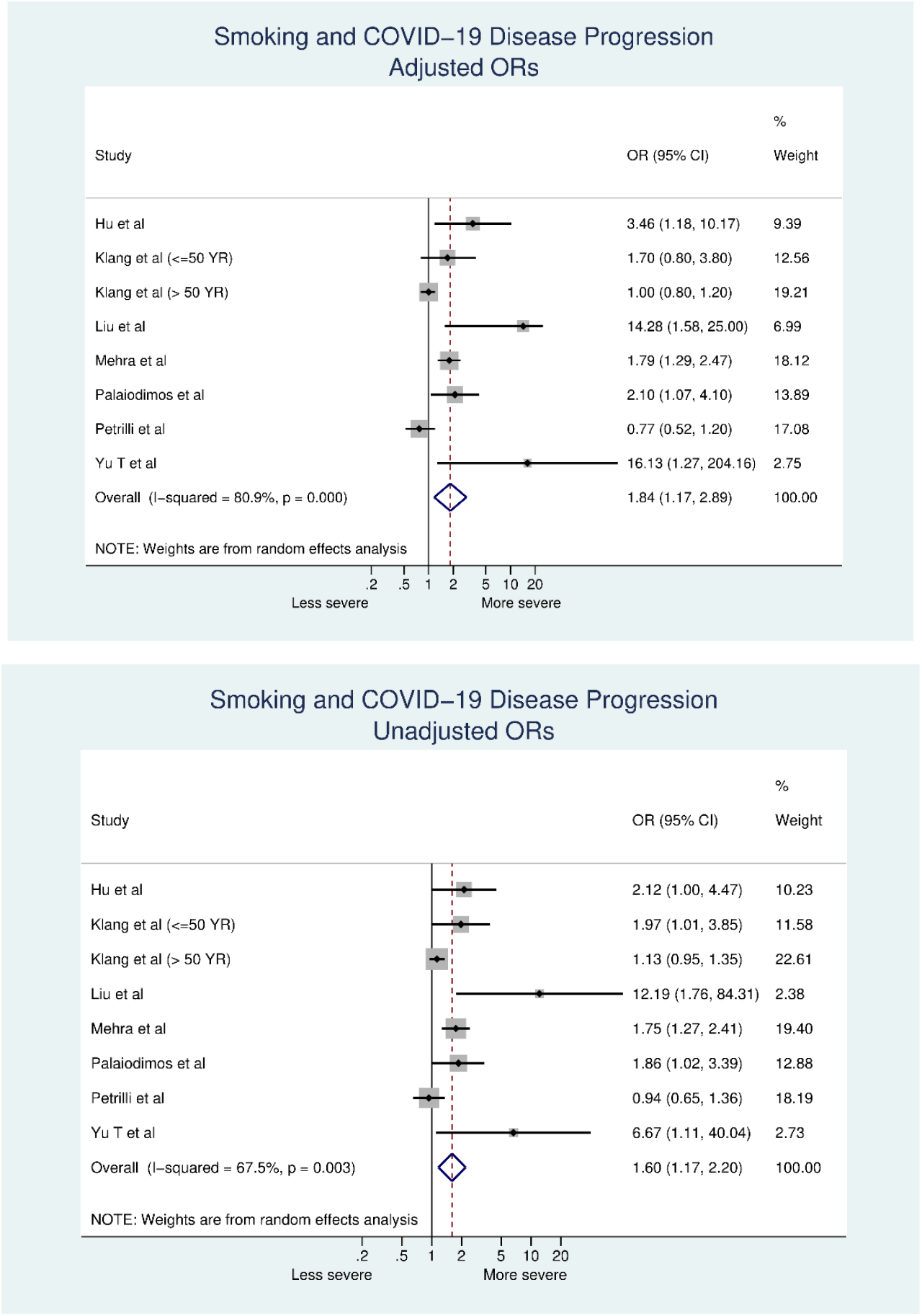
Smoking and disease progression in models that adjusted (top) and did not adjust (bottom) ORs for confounding variables.

